# Technology in Palliative Care (TIP): the identification of digital priorities for palliative care research using a modified Delphi method

**DOI:** 10.1101/2021.06.24.21259307

**Authors:** Amara Callistus Nwosu, Tamsin McGlinchey, Justin Sanders, Sarah Stanley, Jennifer Palfrey, Patrick Lubbers, Laura Chapman, Anne Finucane, Stephen Mason

**Author notes:** Corresponding author Amara Callistus Nwosu, Lancaster Medical School, Lancaster University, Lancaster, LA1 4YG, United Kingdom.

## Abstract

**Background:** Developments in digital health (describing technologies which use computing platforms, connectivity, software, and sensors for health care and related purposes) has the potential to transform the delivery of health and social care to help citizens manage their own health. Currently, we lack consensus about digital health research priorities in palliative care and lack theories about how these technologies might improve care outcomes. Global palliative care need is expected to increase due to the consequences of an ageing population; therefore, it is important for healthcare leaders to identify innovations to ensure that an increasingly frail population have appropriate access to palliative care services. Consequently, it is important to articulate research priorities as the first step to determine how we should allocate finite resources to a field saturated with rapidly developing innovations.

**Aims:** To identify research priority areas for digital health in palliative care.

**Methods:** We selected the digital health trends, most relevant to palliative care, from a list of emerging trends reported by the ‘Future Today Institute’. We conducted a modified Delphi process and consensus meeting with palliative care experts to identify research priorities. We used the views of public representatives to gain their perspectives of the agreed priorities.

**Results:** One hundred and three experts (representing 11 countries) participated in the 1st Delphi round. Fifty-five participated in the 2^nd^ round (53% of 1^st^ round). Eleven experts attended the final consensus meeting. We identified 16 priorities areas, which were summarised into eight themes. These themes were: big data, mobile devices, telehealth and telemedicine, virtual reality, artificial intelligence, the smart home, biotechnology and digital legacy.

**Conclusions:** The identified priorities in this paper represent a wide range of important emerging areas in field of digital health, personalised medicine, and data science. Human-centred design and robust governance systems should be considered in future research. It is important that the risks of using these technologies in palliative care are properly addressed to ensure that these tools are used meaningfully, wisely and safely and do not cause unintentional harm.

## BACKGROUND

Developments in digital health (describing technologies which use computing platforms, connectivity, software, and sensors for health care and related purposes) has the potential to transform the delivery of health and social care to help citizens manage their own health.^1-3^ Currently, we lack consensus about digital health research priorities in palliative care and lack theories about how these technologies might improve care outcomes. Therefore, it is important to articulate research priorities as the first step to determine how we should allocate finite resources to a field saturated with rapidly developing innovations. Global palliative care need is expected to increase due to the consequences of an ageing population; therefore, it is important for healthcare leaders to identify innovations to ensure that an increasingly frail population have appropriate access to palliative care services.^4^ Research demonstrates that, when used well, digital health initiatives improve healthcare delivery and access,^5-15^ and the World Health Organisation (WHO) promotes that digital health should be an integral part of health priorities as a means to improve health on a global scale.^16 17^ To date, many barriers have prevented the meaningful use of digital health in palliative care;^18^ these barriers include expense, inter-operability issues, data privacy and security concerns, lack of effectiveness, equity, and the concern that technology will reduce face-to-face consults between patients and clinicians.^19 20^

Strategic forethought (futurism) can help palliative care leaders to recognise emerging trends, to test, plan and use these innovations in practice.^21^ Consequently, this study aims to identify digital health research priorities and to theorize how innovations in emerging technologies can improve palliative care.

## AIM

To identify research priority areas for technology in palliative care.

## METHODS

We used a Delphi process, informed by the guidance on conducting and reporting Delphi studies (Guidance on Conducting and REporting DElphi Studies - CREDES^22^) in palliative care, to establish the opinion of palliative care experts. A Delphi process can be used as a consensus-based, forecasting process, enabling anonymous expert contributions to predict phenomena.^23 24^ We chose to use the Delphi method for its potential to achieve consensus in areas of uncertainty.^25-28^ We conducted two rounds of Delphi questionnaire, followed by a consensus meeting and a public engagement workshop to establish final consensus on research priorities for digital technology in palliative care. Data collection took place between November 2018 and September 2019.

### Identification of technology trends from the Future Today Institute

We selected technology trends most relevant to palliative care from a list of emerging technology trends reported by the Future Today Institute (FTI - https://futuretodayinstitute.com). The FTI is a multi-professional organisation that uses data-driven applied research to develop models that forecast risk and opportunity across several disciplines, which are mapped into technology trends. The 2018 trend list included 225 emerging trends, which were stratified into 19 categories (Appendix: Future Today Institute 2018 Trends list).

### Selection of technology trends for palliative care

We developed criteria to select the FTI trends, based on recommendations from a UK-based policy report, which reported public and professional views on new types of healthcare data.^29^ We developed the following statement to select FTI trends for inclusion: ‘Trends should involve analysis or use data generated by a patient, caregiver or healthcare professional with potential use in palliative care’. Two authors (ACN and TMc) reviewed all 225 FTI trends. We included 95 (42.2.%) of the trends. We then combined and simplified similar trends to reduce the number to 32 (Figure 1 - Flow diagram to outline study process for identifying research priority areas). To ratify the validity of the trends for palliative care, we conducted a focused literature review to identify examples where these technologies have been used in healthcare. We used an Excel spreadsheet to collate this data for reference.

**Figure 1:**
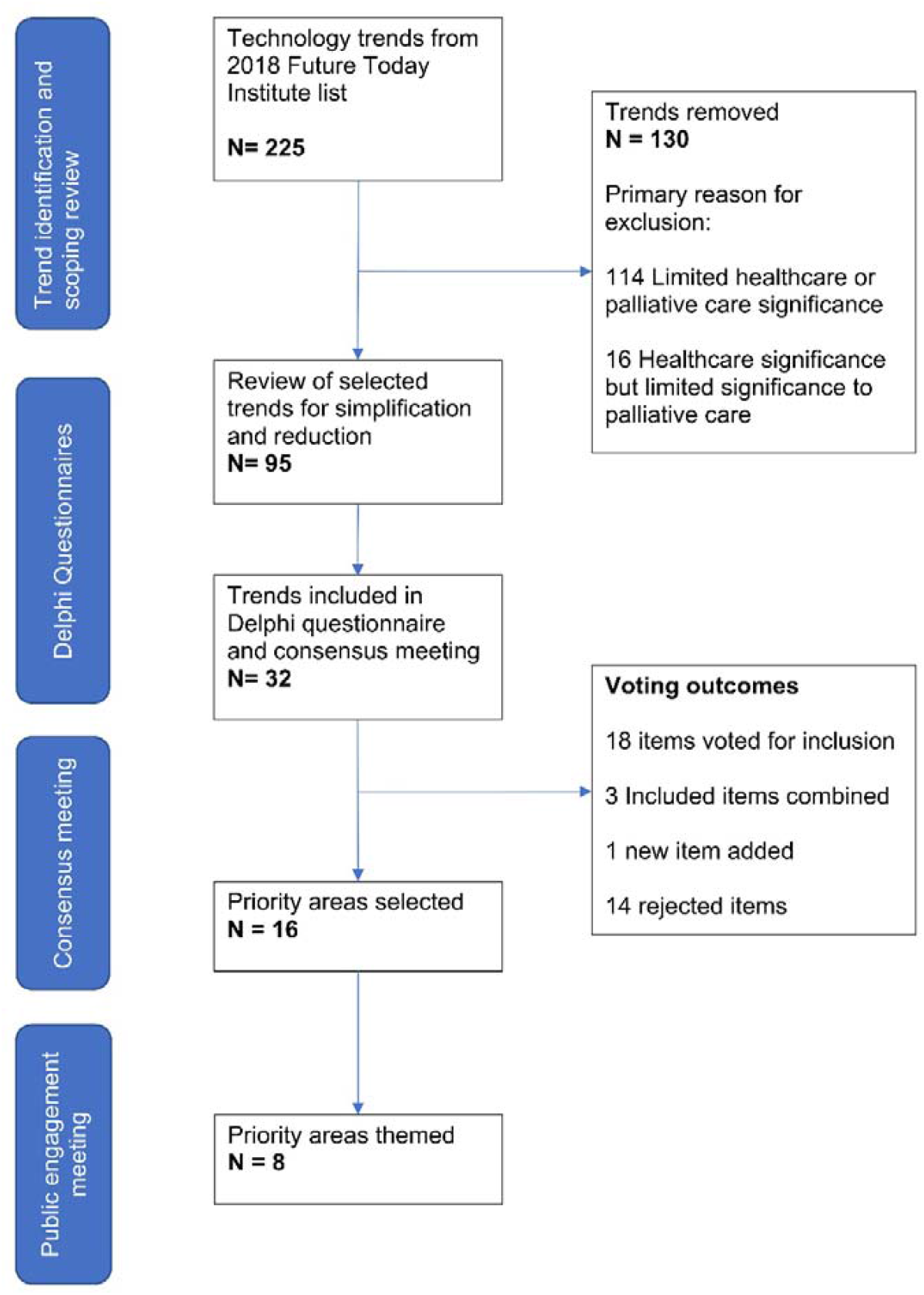
Flow diagram to outline study process for identifying research priority areas.

### Delphi Questionnaire Development

We developed 32 items for inclusion in the Delphi questionnaire, which reflected the 32 trends identified from the Future Today Institute Report (see Figure 1 - Flow diagram to outline study process for identifying research priority areas). We used Google Forms (https://www.google.co.uk/forms/about/) to develop the survey. We designed the questionnaire to collect demographic information (geographic location, age, occupation), and individuals’ rating of importance for each item via a 5-point Likert scale (1 = low priority, 5 = high priority). To ensure that the survey questions were appropriate, we conducted a local pre-study pilot of the questionnaire and supporting materials (Appendix – ‘Delphi Questionnaire’ and ‘Scoping review’).

### Participant Recruitment and Consent

We solicited a convenience sample of palliative care professionals with expressed interest in technological innovation; we used professional networks, social media and email to contact individuals (Appendix - Summary of the networks used to invite palliative care professionals to participate). Consenting participants accessed the study material online to complete an electronic consent form and the 1st round Delphi questionnaire. We invited participants who completed the 1^st^ round questionnaire to participate in the 2nd round.

### Ethical approval

This study was approved by the University of Liverpool Ethics Committee (study approval number 3564).

### Data collection and analysis

Quantitative statistical analyses of participants ratings were undertaken with the statistical software package SPSS 22.0. We used the Interquartile Range (IQR) to determine the level of agreement on the five-point scales for each ‘area’ on the questionnaire. The justification for the levels of agreement were based on thresholds previously used in palliative care Delphi studies, which used a 5-point Likert scale to determine agreement (Appendix: Interquartile Range to be used to guide the Level of Agreement for Delphi responses).^22 30^ We emailed a summary of the 1^st^ round Delphi results to each participant. The email included the following information: (i) a summary of how the participant rated each item in the first Delphi round, and (ii) a summary of all participants’ responses for each item (pooled level of agreement). We provided this information so participants could consider whether they wished to rank items differently in the 2nd Delphi round, based on the ranking data generated by other participants.

### Round 2 Delphi questionnaire

We provided participants with an electronic link to access the 2^nd^ round Delphi questionnaire. We asked participants to answer the same questions that were included in the first-round questionnaire. Participants were required to complete the questionnaire within 4 weeks. We analysed responses from the 2nd questionnaire by IQR to provide a final list of items according to their level of agreement.

### Final consensus meeting and voting

We organised a consensus meeting to agree the trend list as the final stage of the Delphi process.^22^ We invited all participants to attend the meeting at the University of Liverpool, UK. We divided participants into two groups. We attempted to ensure the groups were similar by allocating individuals according to their gender, experience and occupation. We provided participants with the Delphi results, via (i) an oral presentation and (ii) a written summary. ACN and TMc acted as group facilitators and ACN chaired the meeting. We facilitated group discussion and voting. Each item was discussed and debated, and a ‘raised-hand’ vote was undertaken within each group to determine if each item was included or excluded from the final list.

After voting, we compared the outcomes for both groups. We included items if both groups voted for their inclusion. Similarly, we excluded items if both groups voted for exclusion. When the groups disagreed (i.e., one group voting for inclusion and the other voting for exclusion), we facilitated debate with both groups together, which was followed by rounds of voting until consensus was achieved.

### Public engagement workshop

Following the Consensus meeting, we conducted a public engagement workshop with lay representatives to determine their views on the agreed priorities. We used volunteer coordinators from Marie Curie Hospice Liverpool and Liverpool University Hospitals NHS Foundation Trust, to invite palliative care volunteers (by telephone and email).

## RESULTS

### Round 1 *Delphi Questionnaire*

Round 1 included 103 people participants (Table 1 - Demographics of study participants). The median age of participants was 45 years. Most participants were female (n = 65, 63.1%) and had a clinical background (n = 74, 72%). Participants represented 11 countries, most commonly the United Kingdom (n = 88, 85.4%). Most trend items (n = 25, 78%) achieved a median priority rating of 4 or 5 (Appendix: Level of agreement for each ‘priority area’ following both Delphi rounds), which suggested that participants considered most items were important.

**Table 1:**
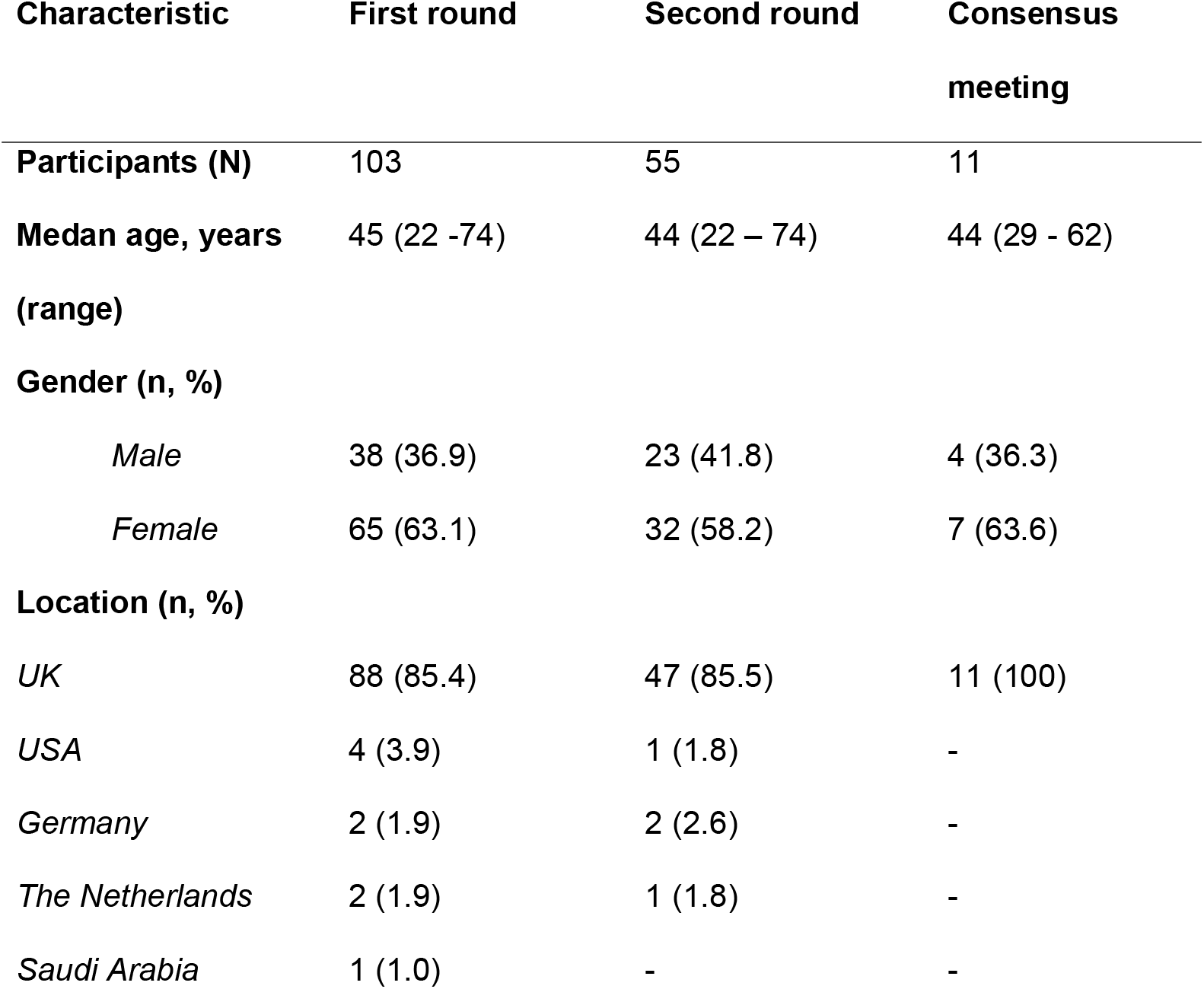
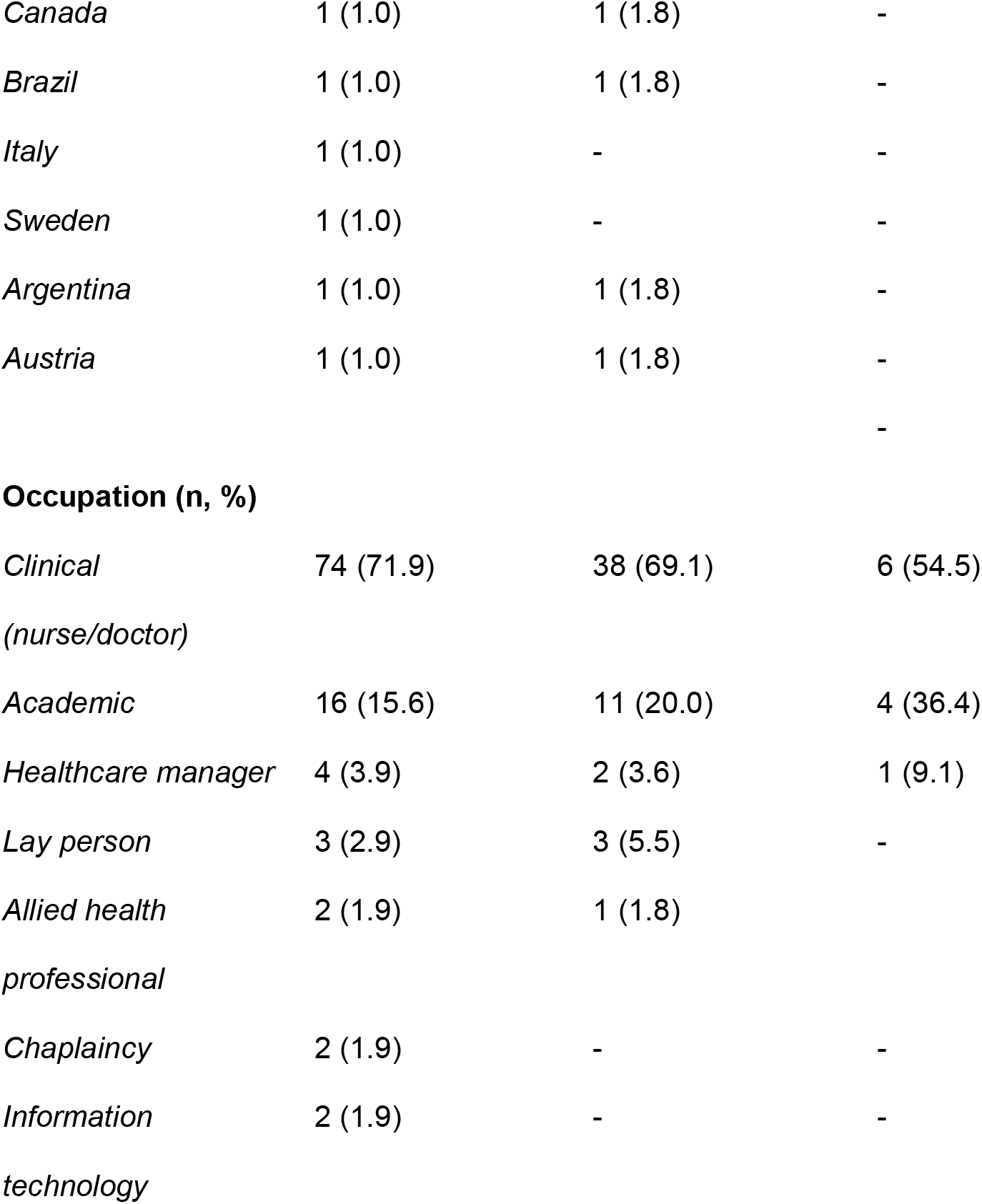
Demographics of study participants.

| **Characteristic** | **First round** | **Second round** | **Consensus meeting** |
| --- | --- | --- | --- |
| **Participants (N)** | 103 | 55 | 11 |
| **Median age, years (range)** | 45 (22 -74) | 44 (22 – 74) | 44 (29 - 62) |
| **Gender (n, %)** |  |  |  |
| *Male* | 38 (36.9) | 23 (41.8) | 4 (36.3) |
| *Female* | 65 (63.1) | 32 (58.2) | 7 (63.6) |
| **Location (n, %)** |  |  |  |
| *UK* | 88 (85.4) | 47 (85.5) | 11 (100) |
| *USA* | 4 (3.9) | 1 (1.8) | - |
| *Germany* | 2 (1.9) | 2 (2.6) | - |
| *The Netherlands* | 2 (1.9) | 1 (1.8) | - |
| *Saudi Arabia* | 1 (1.0) | - | - |
| *Canada* | 1 (1.0) | 1 (1.8) | - |
| *Brazil* | 1 (1.0) | 1 (1.8) | - |
| *Italy* | 1 (1.0) | - | - |
| *Sweden* | 1 (1.0) | - | - |
| *Argentina* | 1 (1.0) | 1 (1.8) | - |
| *Austria* | 1 (1.0) | 1 (1.8) | - |
|  |  |  | - |
| **Occupation (n, %)** |  |  |  |
| *Clinical*<br>*(nurse/doctor)* | 74 (71.9) | 38 (69.1) | 6 (54.5) |
| *Academic* | 16 (15.6) | 11 (20.0) | 4 (36.4) |
| *Healthcare manager* | 4 (3.9) | 2 (3.6) | 1 (9.1) |
| *Lay person* | 3 (2.9) | 3 (5.5) | - |
| *Allied health*<br>*professional* | 2 (1.9) | 1 (1.8) |  |
| *Chaplaincy* | 2 (1.9) | - | - |
| *Information*<br>*technology* | 2 (1.9) | - | - |

### Round 2 Delphi

Fifty-five (53%) of the round 1 participants completed the round 2 questionnaire. The median age was 44 years, which was similar to round 1. More women than men completed the questionnaire (n = 32, 58.2%). The distribution of occupations was similar across both rounds. Fewer countries (n = 8) were represented among the final sample. The final IQR analysis (Appendix: Level of agreement for each ‘priority area’ following both Delphi rounds) demonstrates that most items (n = 21, 65.6%) had *low* levels of agreement, with two (6.3%) and nine (28.1%) items achieving *moderate* and *high* levels of agreement respectively.

### Consensus meeting and final list of priorities

Eleven people participated in the consensus meeting (10.7% of total participants and 20% of second round participants). The median age of participants was 44, and most were female (n = 7, 63.6%). All participants were based in the UK and were mostly from clinical (n = 6, 55%) or academic backgrounds (n = 4, 36%). The debate resulted in agreement, rejection, modification (rewording and combination) of trends, and the addition of a new item, digital legacy (Appendix: Voting outcomes for consensus meeting). We classified the priorities into eight themes which were: big data, mobile devices, telehealth/telemedicine, virtual reality, artificial intelligence, the smart home, biotechnology and digital legacy (Table 2 - Final list of priorities).

**Table 2:** Final list of priorities.

| Theme | Priority | Potential areas for further study identified by public engagement group |
| --- | --- | --- |
| **Telehealth and telemedicine** | a) Use of Telehealth and telemedicine to support patients and caregivers | - How can the telehealth systems be best used to provide remote support for patients and caregivers?<br>- How can video-calling technology be used by health professionals to deliver palliative care? |
| **Artificial intelligence** | b) The use of different AI methodologies (e.g., Machine Learning, Natural Language Processing (NLP), deep learning, neural networks) be used for prediction and screening in palliative care. | - How can algorithms for prediction and screening be developed safely and effectively for palliative care patients?<br>- How can algorithmic driven data be used for palliative care research.<br>- What are the ethical and legal issues concerning use of AI in palliative care?<br>- How can bias be prevented, identified and |
|  |  | addressed? |
|  | c) Ethical and moral issues concerning use of artificial intelligence in palliative care. | - What are the ethical, legal, security and privacy issues of using artificial intelligence palliative care?<br>- How can bias in AI applications be identified and addressed?<br>- Who is responsible for maintaining trust in using AI in palliative care? |
| Big data | d) Collection and use of big data, from Electronic Health Records (EHR) systems. | - How can electronic health records be best designed to optimise use of big data in palliative care?<br>- How can big data be used to improve palliative care on an individual and population health perspective?<br>- What are the training/education needs of staff regarding the use of big data in palliative care. |
|  | e) Governance, data security and regulation of big data use in palliative care. | - What are the responsibilities of stakeholders in the design and use big data, across different aspects of palliative care?<br>- What data security considerations are required for the use of big data in palliative care? |
|  | f) Ethical Challenges of big data health research: | - What are the ethical issues in palliative care research using big data?<br>- What are the implications for informed consent and participation in big data research? |
|  | g) Role of 'big data' and artificial intelligence for palliative care population health management | - How can novel data analysis methods use population level data to support palliative care? |
| **Mobile devices and wearables** | h) Use of mobile devices to support communication, patient monitoring and patient reported outcomes (PROs) | - How can data from mobile devices be used to monitor physical and emotional wellbeing?<br>- How can mobile devices support the collection of patient-reported outcome measures?<br>- How can mobile devices be used to support |
|  |  | communication and information sharing with patients, caregivers and health professionals?<br>- How can mobile devices be used for therapeutic care delivery?<br>- How can advance care planning discussions be best supported, documented and shared. |
|  | i) Development of apps for clinical use in palliative care | - How can apps be designed to ensure safety, efficacy and accuracy?<br>- What are the interoperability considerations of app design/development?<br>- How can risks of app assessment be identified and managed? |
|  | j) Patient-Generated Health Data (PGHD) to promote personalised palliative care | - What data should be collected and what mechanisms should be used for this?<br>- How can sensor-based technologies be best used to support generation of PGHD? |
|  |  | - How can PGHD be used for active and passive palliative care management? |
|  | k) Wearable Health Trackers for Physical Activity<br>Change Detection (PACD) | - How can wearable health trackers support physical activity for people with palliative care needs?<br>- Can wearable technologies be used to detect physical decline in serious illness?<br>- Can wearable technologies help self-management of palliative illness? |
| **Virtual Reality (VR)** | l) Use of virtual reality (VR) for symptom management in palliative care | - How can VR be used for symptom management in palliative care?<br>- What VR equipment, processes and systems offer the best experience for users? |
| **The smart home** | m) Use of Smart Home technologies (e.g., Internet of Things) and sensors for monitoring of health status | - How can the Internet of Things technologies be used to provide palliative care at home?<br>- How can smart (home) assistants support |
|  |  | palliative care delivery?<br>- What are the privacy, ethical and legal issues related to the smart home in palliative care? |
| **Biotechnology** | n) Genome profiling and Personalised Medicine | - How can personalised medicine to improve symptom management or disease specific management in palliative care? |
|  | o) Genetic editing and biomarker technology for earlier disease detection and possible disease management/prevention | - What palliative care complications could potentially benefit from early detection or prevention (e.g., to predict individuals susceptible to development of metastases, pathological fracture or hypercalcaemia)?<br>- Can genetic editing be used to improve management for palliative care? |
| **Digital legacy** | p) Use of technologies which contribute to digital legacy in palliative care | - How can different forms of digital material be used actively to support patients and caregivers to create a 'digital legacy'? |
|  |  | - How should digital legacy be managed after death?<br>- What are the potential risks and ethical issues related to digital legacy? |

### Public Engagement Event

We conducted the public engagement event at Marie Curie Hospice Liverpool, UK, which was attended by six lay representatives, two staff members (nurse and doctor) and a medical student. In this meeting, we presented the Delphi outcomes and we facilitated round table discussions to explore attendees’ views on the agreed priorities. We asked attendees to identify areas within these themes that they wanted researchers to study further.

Our public representatives recommended that future research should: (1) ensure a human centre co-design approach to ensure that technologies are designed according to the needs of individuals and (2) that appropriate governance processes should be in place to evaluate efficacy, effectiveness and ethical issues of current and future digital health tools and systems.

## DISCUSSION

### Summary of main findings

This is first study to identify digital health research priorities for palliative care and provides guidance for researchers, funders and policy makers to consider areas for future research and development. We identified 16 research priority areas for technology in palliative care, representing 8 themes of big data, mobile devices, telehealth, virtual reality, artificial intelligence, the smart home, biotechnology and digital legacy.

### Contribution and strengths of this paper

The outcomes of our detailed analysis (involving a modified Delphi process and patient engagement workshop) indicates further digital health research is needed to study how technology can be best used to support palliative care. Our paper is the first priority-setting paper for palliative care digital health and provides a foundation for digital health focused palliative care research.

### Telehealth and telemedicine

Prior to the novel coronavirus disease 2019 (COVID19) pandemic, researchers highlighted the potential to use telehealth (i.e., technology to support remote clinical access), and telemedicine (i.e., technology to support remote clinical care delivery) in palliative care. These technologies are increasingly used in palliative care;^31 32^ however, many are unevaluated for use in real-world settings.^19 33^ Beyond the pandemic, researchers can consider how these technologies can improve palliative care access (e.g. for remote communities, hard to reach groups) to support new models of care (e.g. tele-palliative care clinics). It is also important to consider barriers (e.g. equity of access, privacy and security considerations) facilitators (ease of use, incentives) and use-cases (e.g. reasons for use) for adoption of telehealth and telemedicine in palliative care.

### Artificial Intelligence

Artificial intelligence (AI) is often used as an umbrella term to describe a number of processes (e.g. Machine Learning, Natural Language Processing (NLP), deep learning, neural networks).^34^ Clinicians and researchers are increasingly using AI to predict survival,^35-38^ classify pain severity,^39 40^ identify quality indicators,^41 42^ and to identify serious illness conversations from electronic healthcare records.^43^ However, most of these studies are exploratory and do not provide recommendations for clinical practice.^18^ Therefore, researchers should explore how different AI techniques can support palliative care research and practice, with consideration to the ethical issues associated with these methods.

### Big data

Big data describes the large amounts of (previously unmanageable) data, which can now be processed by modern-day computer analysis techniques. The opportunities to use routine data to support palliative care decisions for populations and individuals has previously been reported.^44 45^ Currently, there is no consensus for how non-traditional sources of big-data can be meaningfully used in palliative care. For example, there is potential to use patient-generated data (e.g., from wearables) for quality-of-life assessments. Furthermore, open source genomic databases may provide opportunities to study relationships between genetics and health, to inform how data can be used for disease management. Social media, and other forms of online data, are increasingly used to support public and professional communication, and to gain insight on the public attitudes to palliative care.^46-49^ Consequently, researchers should identify what data to collect, and how to best use both traditional and non-traditional sources of palliative care big data.^18 50 51^

### Mobile devices and wearables

Many studies have described how mobile devices and wearables can support palliative care (e.g. remote monitoring of physical activity and symptoms, to deliver wellbeing activity, for documentation of advance care planning, education access/delivery and guideline access).^52-56^ The capability of these devices to collect and store data are increasing; therefore, it is important to determine how this data can be meaningfully used.^57 58^ Researchers have previously described how patient-reported outcomes (PRO) can improve palliative care patients,^59-61^ however, further work is needed to explore how this technology can best support PRO collection (and use) in real world settings.^62 63^ It is important to examine how mobile devices are designed to meet the requirements of palliative care users.^64^ Furthermore, studies should provide more information of how mobile devices can help patients to record their care preferences (e.g., advance care planning).^65 66^

### Virtual reality

Virtual reality (VR) is a human-computer interface technology that uses visual graphics, sounds and other sensory input to create an interactive computer world.^67^ Previous studies have described the potential to use VR to support psycho-social symptoms and wellbeing; however, most work is unevaluated so further research is needed.^68-71^ We recognise the potential of VR to support palliative care education;^72 73^ however, the Consensus group did not identify this as a current priority. Following our study, we recognize that the COVID19 pandemic has accelerated the use of virtual learning environments for medical education,^74^ particularly with the potential to use VR for communication skills training.^72^ Consequently, it is possible that VR for education would rate higher as a priority if this study were repeated.

### The smart home

A smart home describes a living environment where sensor-based systems and internet-connected devices (the Internet of Things) are used for remote monitoring and automation of appliances, such as lighting and heating.^75^ Previous studies illustrate how various technologies can support care for people experiencing decline in their physical function (e.g. virtual assistants and supportive robotics), which highlights the wider role these technologies may have in practice.^76^ Consequently, future work should explore the usefulness of smart home technologies in supporting physical function, and the legal, privacy and ethical issues associated with these developments.^51 3 65 75-77^

### Biotechnology

Biotechnology involves the combination of technology with living things.^78^ Palliative care related developments include use biomarkers to predict survival,^79 80^ constipation, ^81^delirium,^82 83^ and the personalisation of cancer pain according to genetics.^84-86^ Consequently, it is possible to imagine future scenarios where technologies are used for early identification (and prediction) of clinical issues, facilitating personalised treatment for the individual (e.g. early identification and management of pathological fracture).

### Digital legacy

A digital legacy is the digital information available about someone after death, such as social media, photos, videos and gaming profiles.^87^ The volume of digital information generated by citizens is increasing, which creates new challenges after death.^88^ The increasing use of cloud storage and social media is contributing to uncertainty of data ownership, which creates difficulties for caregivers to manage the digital legacy of the deceased. Studies demonstrate that healthcare professionals can positively support their patients to manage their digital legacy.^87 89 90^ However, digital legacy is not routinely discussed in clinical practice, which means that we generally do not know how individuals want their data to be managed after death.^91^ Therefore, we believe that researchers should explore how patients and caregivers can be supported to manage their digital legacy after death, with exploration on the different methods and materials that can be used.

### Relation to previous work in this area and areas of interest following the novel COVID19 pandemic

Our study is synergistic with previous work, which has been conducted across the theme areas.^19 33^ We acknowledge that our study pre-dates the pandemic and it is possible that the priorities we identified may now have shifted. However, we believe our research findings are valid as the digital health innovations adopted during the pandemic were in sync with our priority list. (Appendix: Examples of technologies used in palliative care during the COVID19 pandemic).^32 33^ Telehealth was commonly used during the pandemic, with many palliative care services using this to provide remote clinical support,^92-104^ to communicate^105^ and for education.^106^ Technologies were used to maintain connection, and to develop communities of palliative care practice.^107 108^ VR was used to provide psychological care and symptom management.^109 110^ In general, the findings these studies describe potential benefits of digital health; however, the rapid implementation of these technologies has created a number of challenges (e.g., technical issues, data security and wellbeing considerations) which, require further evaluation.^105^

### Limitations

It is possible that recent developments were not reflected in the priority list due to ongoing advancement of healthcare technologies. For example, the FTI trends list is now in its 2021 version and includes new trends such as, home medical laboratory tests and remote metabolic monitoring. Therefore, it is possible that relevant areas are absent from this analysis. Also, a weakness of digital health research is the rapid change associated with technology, which may cause the findings of this study to lose relevancy over time.

Our decision to reduce the number of trends from 95 to 32 items, has broadened the focus of the list, which means it is possible that more specific and technical areas were not explored in greater depth (e.g., faceprints, voiceprints, chatbots etc etc). It is also possible that our Delphi participants will have different views on priority of some areas post COVID19, due to the observed increase of digital health in practice. Questionnaires were mostly completed by participants arising from English-speaking countries, meaning that the experience of non-English speaking populations may not be reflected. It is possible, due to the novel nature of some areas, that participants gave more priority to familiar areas and therefore, less priority to unfamiliar areas. The final priority list may not represent non-UK healthcare systems, as the consensus meeting was only attended by UK residents. Furthermore, people from different professional backgrounds (including cultures and settings) may assign different levels of priority to trends, due their experience, work-requirements and personal beliefs. As most participants were clinically-focused, it is possible that the priorities were orientated to clinical-utility, rather than methodology.

### Relevance to research, practice and policy

Decision-makers should ensure that technology is relevant to the needs of the palliative care user, as these requirements will influence the design, use and function of systems. For example, healthcare professionals may generally use technology to access patient data and communicate with other professionals, whereas patients may wish to access their own health data and to contact healthcare services. Further research is needed to develop specific use-cases for these scenarios, to ensure that the technology can be used meaningfully to achieve the intended outcomes. Furthermore, as the user requirements of people with palliative care needs may differ from the general population^111^ and because we lack resources for wide-spread implementation of all technologies currently, it is important that digital health studies provide the data needed for determine best practice, and to help identify the barriers and facilitators for adoption.

Researchers should use appropriate methodologies to explore these questions and should also study associated areas, such as ethical issues, data security, and design. It is important that researchers work with the public, as the comments of the lay representatives in our study (from both the consensus meeting and the public engagement workshop) described concerns about the use of personal data. Policymakers should consider issues related to governance and ethics of current, and future, digital systems. From a design perspective, we suggest that palliative care professionals work collaboratively with creative industries (e.g., designers, developers and engineers) to ensure that designed technologies fulfil the user requirements for specific palliative care use-cases.

## Conclusion

The identified priorities in this paper represent a wide range of important emerging areas in the field of digital health, personalised medicine, and data science. Human-centred design and robust governance systems should be considered in future research. Transdisciplinary studies using appropriate methodologies are required to further study this priority list. It is important that the risks of using these technologies in palliative care are properly addressed to ensure that these tools are used meaningfully, wisely and safely and do not cause unintentional harm.

## Supporting information

Scoping Review - all sections

Summary of networks used to invite palliative care professionals to participate

Voting outcomes for consensus meeting

CREDES checklist TIP study paper 2021

Delphi questionnaire - Google form

Examples of technologies used in palliative care during the COVID19 pandemic

Future Today Institute Trends 225 list

Interquartile Range to be used to guide the Level of Agreement for Delphi

Level of agreement for priority areas both Delphi rounds

## Data Availability

All study data are available from teh manuscript and supplementary files.

## ACKNOWLEDMENTS

This research was supported by the following funding streams: Liverpool Clinical Commissioning Group (CCG) Research Capability Funding (researcher salary), £20,970. Wellcome Trust, Public Engagement Grants Scheme (Public Engagement Event costs), £360. Liverpool Clinical Commissioning Group (consensus meeting costs) £807. The posts of ACN and SS were funded by Marie Curie: https://www.mariecurie.org.uk/

## AUTHOR CONTRIBUTIONS

The author’s responsibilities were as follows:

Study design: ACN, TMc

Data collection: ACN, TMc

Paper writing: ACN, TMc, JS, JP, LC, SS

Critique and review of the final manuscript: ACN, TMc, SS, LC, JP, AF, PL, JS, SM

## DATA AVAILABILITY

The authors declare that the data supporting the findings of this study are available within the paper and its supplementary information files.

## LIST OF ABBREVIATIONS USED

AI: Artificial Intelligence
EHR: Electronic Health Records
ePROM: electronic patient-reported measures
FTI: Future Today Institute
ML: Machine learning
NHS: National Health Service
NLP: Natural Language Processing
PACD: Physical Activity Change Detection
PGHD: Patient-Generated Health Data
PROMs: Patient reported outcome measures
VR: Virtual Reality

## APPENDICES

- Future Today Institute 2018 Trends List.
- Technology in Palliative Care study scoping review.
- Delphi questionnaire (Google forms).
- Voting outcomes for consensus meeting.
- Summary of the networks used to invite palliative care professionals to participate.
- Interquartile Range to be used to guide the level of agreement for Delphi responses.
- Level of agreement for each ‘priority area’ following both Delphi rounds.
- Examples of technologies used in palliative care during the COVID19 pandemic.

