## Supplementary material for "Technology in Palliative Care (TIP): the identification of digital priorities for palliative care research using a modified Delphi method": Scoping Review - all sections

**Technology in Palliative Care (TIP) Study: Priority Setting to Improve the Care of Patients with Advanced Cancer.**

**Scoping Review: examples, uses and developments.**

The development of digital technology within healthcare presents a new and exciting opportunity in the way that healthcare is designed, delivered, and experienced. Purposeful development, considered use and wider adoption of technology has the potential to transform and drive improvements in the quality and efficiency of healthcare provision and patient experience (Honeyman et al 2016). This provides an opportunity for services to optimise 'patient centred care' by reforming the way patients engage with services, as well as supporting people to manage their health and wellbeing (Imison et al 2016, Ham et al 2012).

Although digital technology may appear to be a new concept within healthcare its development and uses can be mapped since the 1960's, with the first use of digital technology focussed on administrative, financial and research purposes. It wasn't until 1991 however, that the first National IT Strategy for the NHS was published, and the 2000's before any real significant investment in digital technology was recommended and actioned (Kings Fund https://www.kingsfund.org.uk/audio-video/digital-timeline). Despite this slow evolution, the development of digital technology has become increasingly prominent within health care. In the last 5 years there has been an increasing policy focus on the digitisation of healthcare, with the most recent Watcher review (*Using information technology to improve the NHS*, 2016) recommending current IT systems are revolutionised, with a focus on 'paperless' systems and digital technology, and an acknowledgement of the need to harness the enormous potential to really make it work for the NHS.

With this renewed investment and policy drive, comes an opportunity to consider the way forward for digital technology in healthcare, specifically for patients with cancer. This project will look to set objectives (or 'priority areas') for the future development of data-driven healthcare applications, to improve the care of people living with advanced cancer.

In order to aid the process a scoping review of both the literature and internet has been undertaken in order to identify examples of how digital technology has been used to support healthcare, including identifying those with a focus on advanced cancer. This scoping review used themes identified by the Future Institute Today (FIT) (*2018 Tech Trends Report*), focussing on the 'Health Technologies and Wearables' and 'Smart Homes and Internet of Things' sections within the report, as a basis for the search. These themes were then used to structure the review, to identify data-driven healthcare applications currently in use or in development, and to highlight where these technologies have a palliative care or advanced cancer focus. These applications will be categorised and 'listed' for inclusion on the round 1 Delphi questionnaire. This questionnaire will be designed to enable participants to rank the order of importance of identified themes/uses of data-driven healthcare applications.

**PROCESS:**

The scoping review used a 6 stage methodological framework (Levac et al 2010), to add structure to the review, and to aid reporting for clarity of process:

1. **Identifying the research question**

For this study, the 'research question' is to explore and categorise the examples, uses and developments in data-driven technology for patients with palliative care needs.

2. **Search strategy: relevant 'data-driven technologies'**

Key concepts, or 'themes', were used to structure the literature database and the internet searches. It is important to keep the scoping exercise broad, however it is also important to define and articulate the scope of enquiry (Levac et al 2010). The FIT 2018 Tech Trends Report was used as the basis for the review, to identify key concepts/themes:

- Health Technologies and Wearables
- Smart Homes and Internet of Things
- Machine learning/artificial intelligence
- Virtual reality and augmented reality
- Robotic technology

3. **Criteria for selection of 'data-driven technologies'**

Relevant data-driven technologies were identified using the criteria below, and screening was undertaken to eliminate duplicates. Specific data-driven technologies were then selected based on the following criteria:

Inclusion criteria

- Is a technology that uses data to gather/store/report/analyse patient data
- Has been used to support healthcare
- Is in English language.

Exclusion criteria

- Is not a technology that can be used to gather/store/report/analyse patient data
- Has not been used to support healthcare
- Is not in English language.

4. **Charting the data: extraction of a priori information**

In order to create a comprehensive database of the use of digital technologies in healthcare, the following information was collected from each identified 'technology':

- Description of the technology/area of development
- Target population
- Advanced cancer focus
- Reference to relevant article/website

**5. Collating, summarizing, and reporting the results**

Key information listed above will be collected and stored in an Excel spreadsheet, against each identified data-driven technology/area of development. The end result will be the development of a repository which will provide an index of the range of data-driven technologies that have the potential to support the delivery and experience of healthcare, and specifically palliative care. Table 1 below presents the results of the scoping review.

**6. Consulting with stakeholders to inform or validate study finding:**

This database will be, as far as we know, the first to identify and categorise the use of digital technology for use in patients with advanced cancer. Information gathered from this scoping review will be used to inform the development of the 1st round questionnaire for the Delphi study to be conducted, to identify the 'top 10 priority areas' for future research into data-sharing technology in palliative care.

**Table 1: Scoping Review Results**

| Theme | Category |
|---|---|
| **'Big Data'** | **Examples/development** |
| | Big Data: "health-related data – including health history, symptoms, biometric data, treatment history, lifestyle choices, and other information-created, recorded, gathered, or inferred by or from patients or their designees (i.e. care partners or those who assist them) to help address a health concern" (Shapiro et al., 2012).<br><br>**1.1 Development of Electronic Health Records (EHR) and systems based approaches to collection and utilisation of 'big data':**<br>Digitisation of all aspects of clinical care and documentation has the potential to provide important information about a patient's condition. Although still emerging, this has prompted investment into the development of strategies to incorporate data from these devices, as well as from EHR and other digital systems, to enhance the information available about a patient and their condition. This has the potential to augment healthcare provider's knowledge and understanding of patients and inform clinical care.<br><br>Examples:<br>• Leading companies in the generation of EHR: GE Healthcare; Meditech; Allscripts; eClinicalWorks; Cerner; Validic; HumanAPI; Apple; IBM; Microsoft; Qualcom; Tacito; Alphabet; Medicare; Medicaide; national health systems; insurance companies<br>• Smart phones, electronic health files, fitness trackers<br><br>**1.2 Patient-Generated Health Data (PGHD) and 'big data' – what to collect and how: harnessing and using 'physical function' data via sensor-based technologies**<br>PGHD – the quantified Self: The potential to capture clinically relevant data outside traditional care settings, by patients: Commercially available physical activity monitors are being rapidly adopted by patients and researchers and have the potential to greatly improve our ability to quantify physical activity in patients with cancer (Beg et al 2017).<br><br>The increased availability of PGHD has the potential to generate mass amounts of clinically useful data. Ethical considerations have become apparent however, such as the implications for using 'patient owned' data – data generated by the patient rather than routinely collected as part of the patients 'usual care' – for healthcare monitoring. The ever increasing development and use of data-driven technology in healthcare makes consideration of these issues an imperative. |

Potential benefits of this technology include: deeper insight into a patient's condition; more accurate patient information, particularly when of clinical relevance; insight into a patient's health between clinic visits, enabling revision of care plans for improved health goal achievement, while avoiding unnecessary clinic visits.

Examples:

- Apple - "HealthKit" (iOS8) (partnerships with Mayo Clinic/the EPIC electronic health): records data captured via sensors which could then be uploaded into electronic health records and (at some point) be available on an iPhone https://www.forbes.com/sites/danmunro/2014/06/03/apple-gives-epic-and-mayo-bear-hug-with-healthkit/#4cc1f80f6bb3
- Contribution to research and population level data: potential to help researchers gather large amounts of data on health problems and their determinants using their smartphones etc (Kings Fund 2016, Cohen et al 2016)
- Methodological research around feasibility and modelling of PGHD streams is needed: Research to identify which types of PGHD are the most useful for improving health outcomes and quality of care both on an individual and population level (Wood et al 2015).

**1.3Governance in the use of 'Big Data' – regulation and responsibility**

With the development of technology comes an increase in the levels and type of data that is collected, stored, transferred and utilised. This poses a unique challenge for healthcare, particularly with regards to patient confidentiality. For example, who has 'responsibility' for ensuring privacy, confidentiality, or data security?

Examples:

- Privacy risks and 'wellness' apps – huge variation in privacy practices observed in clinically-accredited health and wellness apps which has highlighted the responsibilities of regulators. Establishing standards for accreditation processes as part of the development of these apps is a paramount, as is monitoring these standards and having protocols to intervene if 'accredited' programmes or apps do not manage these risks effectively. If patients or the public are deterred from using apps because of questions of trust, then the potential clinical benefits of mobile health will not be realised (Huckvale et al 2015).
- Machine Learning tools – System security through user and entity behaviour analytics has the potential to increase security of data collection, transfer and storage however this is still an emerging field.
- Investment is required to develop practice workflows and protocols related to PGHD collection and use; data storage, accessibility at the point of care, and privacy concerns; ease of using PGHD data (Cohen et al 2016).
- Optimising development and use: design, infrastructure and IT support:

- Paradigm of ‘Preventive, Personalized, and Precision Medicine’: Seamless use of technology and infrastructure support is a global challenge for the provision of quality healthcare at affordable cost for patients and healthcare providers at point-of-care (POC) locations including homes, semi or pre-clinical facilities, and hospitals (Dhawan 2016)

**1.4 Cybersecurity and ransomware – Packaging/capturing data and keeping it safe/transfer of information**
With developments in electronic patient records and data-sharing mechanisms, data security is an important issue. There is a need to maximise the security of this information particularly when it is shared, disseminated and transferred, for example, between organisations or across a network of computers within large care providing organisations. A huge area of development in recent times has been the development of block chain technology: A secure process of sharing information between open and private networks. In essence the blockchain is used to verify ‘transactions’ as they transfer along the ‘chain’ – benefits to the safety of healthcare information means that confidential information can only be shared with ‘verified’ personnel.

Examples:

- Development of Block Chain technology (Mertz 2018, Liu et al 2017) – potential to generate vast amount of patient data, meta-data, which could be used to transfer information between healthcare professionals and healthcare services seamlessly and securely, as well as provide opportunities for research and population health management. Too early to know exactly how this technology will be used and leveraged along the way (Mertz 2018):
    - Capital expense and cost of operation
    - Immense amount of clinical data – decisions about what is ‘on chain’ or ‘off chain’ imperative
    - Integration of healthcare regulation and the technology revolution required – ensure that ‘data silos’ are avoided and there is opportunity for greater sharing of information

**1.5 Ethical Challenges in Big Data Health Research: implications for informed consent and participation**
Pervasive capturing and analysis of diverse data types fuel new capabilities in health research. As a result, data-driven discoveries are expected to play an increasingly important role in biomedical research. The huge variety of ‘big data’ opens the door to previously unimaginable applications of data mining for disease prevention, diagnosis, and treatment. There is a potential for vast amounts of data to be available, whether it is through PGHD or EHR’s or even social media and other non-traditional health related data, raising important ethical challenges with regards to how patient data is used (Ienca et al 2018). Increased collection, storage, and use of data within healthcare organisations, and shared across partners, requires scrutiny and principles of good practice.

- The use of ‘Big Data’ offers a unique opportunity to identify palliative care needs, as well as inform population and

| | |
|---|---|
| | individual-level interventions. Future research is essential to address the challenges faced by Big Data analysis in palliative care (Nwosu et al 2018).<br>• Use of Big Data from publically available sources does not require research ethics but data from private sources raises ethical issues (Rothstein 2015)<br>• Concept of 'systemic oversight' in biomedical big data research. This approach is based on six defining features (adaptivity, flexibility, monitoring, responsiveness, reflexivity, and inclusiveness) and aims at creating a common ground across the 'oversight pipeline' of biomedical big data research – including enhancing granularity of informed consent and specifying legal provisions to address informational privacy and discrimination concerns in data-driven health research although more work needed to understand this (Vayena and Blasimme 2018).<br>• The ethics of Big Data is an innovative and emerging field which lags behind the exponential level of growth of technological development. Big Data has the potential to generate "destabilising amounts of information, which can challenge accepted social and ethical norms" (Mittelstadt 2016).<br>• Cambridge Analytica scandal – should be a galvanising force for research ethicists and increased research scrutiny over the use of Big Data (Editorial, Nature 2018) |
| **Wearable Technology** | **Examples/development** |
| | **2.1 Wearable's and Sensor-based technologies: Assessment of physical function and assessment/amelioration/monitoring of physical symptoms**<br>(Future Institute Today tracker shows 538 wearable devices designed to use sensor based technologies)<br><br>Increased development of wearable technology has the potential to revolutionise healthcare:<br>• low-cost solutions for ubiquitous, all-day, unobtrusive personal health monitoring<br>• enable early detection and better treatment of various medical conditions as well as disease prevention<br>• better understanding and self-management of chronic diseases<br>Further investment in research and development has been suggested to fully explore these potentials including applicability, relevance, ease of use and reliability of developed wearable technologies (Pantelopoulos and Bourbakis 2010), particularly their impact on patients with advanced cancer (Beg et al 2017, Gresham 2018).<br><br>Examples:<br>• Smart Thread (FIT 2018): Smart Threads – early development experimental stages but with positive results as a potential diagnostic or monitoring, or as a mechanism for displaying information:<br>○ Tufts University – nano-scale sensors in surgical thread to measure physical (strain and temperature) and chemical (pH and glucose) markers (Mostafalu et al 2016); |

| | |
|---|---|
| | - University of California Berkely School of Information – Smart Threads that change colour, turning fabric into a computerised 'Dynamic Display' (Devendorf 2016).<br>• Physical Activity Change Detection (PACD):<br>- Fitbit Charge Heart Rate® devices used with two separate Alzheimer's disease populations. Data was continuously recorded to evaluate PACD and has shown promising results to detect physical decline (Sprint et al 2017)<br>- 'Online Behavioural Change Detection' system using sensor based tech and machine learning – activity recognition algorithms have been designed for wearable, phone, home, video, and other sensors using machine learning techniques. Feasibility of the 'Eldercare Monitoring System' (EMS) has shown promise in detecting behavioural changes, to identify triggers for early intervention by caregivers (Conference paper: Tam et al 2016).<br>• Reducing burdensome interventions for patients: optimising monitoring and recording of physiological signals:<br>- Sleep/wake analysis – sleep data collected through wearable technology means fewer requirements for physical monitoring of patients overnight (Fu et al 2017).<br>- Unobtrusive epidermal devices with small-scale integration (such as skin patches etc): electronics with diverse functions, such as electrophysiological, temperature, strain responsive sensors, transistors, photodetectors, radio frequency generators etc, which can provide information on electrical activity produced by the heart, brain and skeletal muscles:<br>▪ Fully integrated wearable sensors for perspiration analysis – although with potential for monitoring a wide range of non-invasive, personalised diagnostic and physiological monitoring applications (Gao et al 2016).<br>▪ Flexible, textile and stretchable electronics are emerging research areas: ultra-lightweight design for imperceptible plastic electronics. Applications to: consumer and mobile electronic appliances, architectural design, robotics, emergency response, sports, healthcare and biomedical systems (Kaltenbrunner 2013)<br>▪ Integration of 'self-healing healthcare devices' (plastic, or 'polymer', with ability to 'heal' itself at body temperature) with smart technology to develop smart wearable devices with desirable long-term stability and durability for future clinical applications (Jin et al 2017).<br>• Wearable Health Trackers:<br>- Apple - Smart Watch<br>- Samsung - Simband (https://eu.usatoday.com/story/tech/personal/2014/05/28/samsung-health-announcement/9669879/)<br>- Wearable activity monitors to assess performance status and predict clinical outcomes in advanced cancer |

patients: Fitbit Charge HR® activity metrics and performance status. Activity data suggested a trend for the prediction of clinically relevant adverse events, 30-day morbidity, and 6-month survival (Gresham et al 2018).

- Wearable fabrics for the detection of stimuli (physical functioning e.g. temperature, motion, strain, activity monitoring) - This functionality presents many possibilities; it highlights 'costume' (clothing) as an "active and activating object capable of dynamically intervening between the body and its environment" (Chittendon 2017). There is the potential for further developments for patients with palliative care needs although it is a developing field:
  - Healthcare monitoring: robust and versatile smart-fabric sensor, for the detection of stimuli such as temperature, motion, strain – physical functioning (Shahadid and Seung-Hwan 2018).
  - sportswear design and fabric science: example of tennis costume design incorporating temperature, sweat and muscle control properties, as well as electronic sensors to capture biometric and biomechanical data whilst actively engaged in activity – 'big data' (Chittendon 2017).
- Body area network (BAN) technology: a system which remotely monitors patient health status using tiny wireless sensors devices placed in or around the human body (Baba et al 2018, Sawaneh et al 2018)
  - Security, privacy and data reliability all areas for further research and development
  - HealthMate Smart Wearable System for Health Monitoring (SWSHM) integrated on the Wireless Body Area Networks (WBAN): body position sensors allow patients to monitor body temperature, electrocardiogram, pulse and oxygen in blood, airflow etc – the HealthMate system then uses AI to examine the data, so that alerts can be sent to health professionals if action is required (Omer et al Al-Salihi 2017).
  - Bioelectrical impedance:
    - BI vector analysis (BIVA) of hydration status in advanced cancer – monitoring hydration needs (Nwosu et al 2013)
    - Integration of hydration monitoring with mobile devices is predicted to be the future of hydration assessment technology (Asogwa and Lai 2017)

**2.2 Wearable technology – privacy and ethical considerations**

The proliferation of healthcare data monitored and collected via wearable technologies has highlighted the need for increased 'privacy solutions' in the use of such technology. However, very little research has been performed from an ethical perspective addressing privacy, privacy policies, informed consent and stakeholders (Segura Anaya et al 2018).

Benefits:

Benefits of wearable technology advertised by developers, for users and patients, are varied and ambitious including:

| | |
|---|---|
| | enhancing quality of life, promoting lifestyle changes and saving time and money in medical appointments.<br><br>Privacy:<br>Wearable devices require different degrees of privacy intrusion to collect data, making it important to consider ethical implications from different stakeholders' perspectives (Spring 2009)<br><br>Security risks:<br>Security and privacy risks – for example hacking attacks such as remotely modifying, reprogramming or disabling the wearable device, causing intentional malfunction and putting patient safety at risk (Camara 2015).<br><br>Issues of informed consent are raised, for example users and patients have the right to decide when to share their personal information however there is a critical gap in this area of research in this area (Spring 2005). |
| **Mobile Devices** | **Examples/development** |
| | Mobile devices have become ubiquitous and integral to the way that we conduct our lives. From laptops, hand held devices, mobile diagnostic and clinical devices to name a few, these advances in technology have altered the way in which healthcare data can be collected as well as the way clinical procedures and patient monitoring can be carried out.<br><br>**3.1 Use of mobile devices to gather patient reported symptom outcomes (PRO's)/Patient Performance Status:**<br>Smart Phones: sensors for health-relevant data (movement, location tracking, touch-screen interface).<br>So far it has been suggested there has been poor utilisation of this potential, e.g. NHS App Library has had little success so far (Huckvale et al 2015).<br><br>Examples:<br>• Smart Watch for pain assessment for patients with advanced cancer (Nwosu et al 2017)<br>• Wearable activity monitors (Fitbit Charge HR®) to assess performance status and predict clinical outcomes in advanced cancer patients: association found between activity metrics and performance status and the association between the wearable activity metrics and survival, the occurrence of serious adverse events, as well as the correlation between the wearable activity metrics and PROs in the domains of pain, physical functioning, and fatigue (Gresham 2018).<br>• "ANAPA®" wearable device to measure patient reported pain scores by patients with advanced cancer: comparing clinician assessment and PRO (https://clinicaltrials.gov/ct2/show/NCT03117075# Clinical Trial – active since 2017 – not yet recruiting). Principal Investigator: YOONHO KO, MD, PhD, Uijeongbu St. Mary's Hospital, the Catholic |

University of Korea.

**3.2 App design: Clinical input – Safety, efficacy, accuracy and assessment of risk**
Apps are often freely available and often unregulated – especially in the case of health care applications. Research is required to develop specific strategies to underpin App development. For example developing working policies that encompass regulatory standards and developer responsibility including ensuring meaningful clinical input, as well as paying attention to the accuracy, usefulness and efficacy of the designed App.

Efforts to assure the quality of medical apps need to be responsive to the changing features and risks posed by apps, for example considering governance as well as clinical factors. Developers need to recognise the differing needs of patients using their apps, for example whether they are using them on their own initiative, or they are using them specifically related to their clinical care.

Examples (highlighting risks):
- Smartphone: Insulin dose calculations shown to be problematic and potentially dangerous for diabetic patients (Huckvale et al 2015)
- Mobile Apps for asthma (Huckvale et al 2015)

**3.3 Portable Hospital-level screening/diagnostics in the home:**
The development of portable devices, such as x-ray machines, blood-testing kits etc to support health care, has the potential for profound consequences for the way we configure our health care system.
Examples:
- 'Hospital in the home' - Dr Michael Montalto (https://www.kingsfund.org.uk/audio-video/dr-michael-montalto-hospital-home-victoria-australia):
  - Aim is to challenge the concept of 'inpatient/outpatient' – devices that can generate reports and aid diagnosis without the need for hospital attendance.
  - Increase efficiency in healthcare provision – however expensive to set up – no standard definition of 'hospital in the home'.
- Point of Care Technologies: Enabling crucial healthcare equipment to be easily transported and efficiently used at the 'point of care' can have huge impacts
  - Flexible substrates: Development and use of ultra-thin chips for high-performance flexible electronics to improve point-of-Care (POC) Diagnostics: combat challenges of paper based POC devices such as pregnancy tests and glucose strips, or inflexible devices – next-generation POC diagnostic tools to meet the need for

| | |
|---|---|
| | patient care and personalised healthcare monitoring in non-hospital and home settings. Potential to be integrated into wearable devices with wireless communications for personalized health monitoring, and ability to be customized for POC diagnostics (Wang et al 2016).<br>○ The global challenge of providing quality healthcare at affordable cost leads to the proposed paradigm of Preventive, Personalized, and Precision Medicine: requires a seamless use of technology and infrastructure support for patients and healthcare providers at point-of-care (POC) locations including homes, semi or pre-clinical facilities, and hospitals (Dhawan 2016). |
| **Telehealth/eHealth** | **Examples/development** |
| | Developments in telehealth/eHealth have potential to enhance and improve palliative care delivery and the experience of patients and their families. Due to the projected increase in complex, ill, and elderly patients with limited mobility, development of this technology has been cited as being increasingly important to the sustainability of the healthcare system (Worster 2017).<br><br>**4.1 Telehealth/eHealth to support patients and their families in their own homes**<br>Telehealth/eHealth has the potential to improve remote communication and contact between patients/carers/clinicians, for example where there may be reduced ability for face to face contact.<br><br>**4.2 Telehealth/eHealth for electronic patient-reported outcomes (ePRO)**<br>Growing evidence demonstrates that electronic patient-reported outcome (ePRO) systems are feasible, well received by both patients and clinicians, and equivalent to original paper-based systems. Further research of ePROs is needed (Klagholz et al 2018).<br>Examples:<br>• Internet communication ‘Clinician Report’: online symptom reporting system for patients with advanced cancer – shown to reduce ‘emotional distress’ of caregivers when given access to ‘Clinician Report’ (Chih et al 2013)<br>• Telemedicine to support outpatient palliative care in advanced cancer (Hennemann-Krause 2015)<br>• eHealth system designed for caregivers that alerts clinicians to worrisome changes in patient health status (Gustafson 2017)<br>• Service providers consider telehealth has the potential to augment current service provision however there is concern that the introduction of these technologies also has the potential to alter the dynamic of relationships between patients and families and community palliative care clinicians (Collier 2016). |

| Virtual Reality (VR) | Examples/development |
| --- | --- |
|  | **5.1 Virtual Reality – Distraction therapy to ameliorate symptoms – pan, distress, anxiety**<br>VR is based on the concept of 'immersion', for example it can be used to distract patients undergoing medical procedures in ways that are not possible with other techniques. Immersion enables the user's attention to be focused on the virtual environment rather than the intervention/procedure.<br><br>Research in VR is constantly expanding, and research shows it has wide ranging applications within healthcare. A wide evidence base of research exists on the effectiveness of VR for reducing acute and chronic pain, as shown in a comprehensive search conducted in 2014 (Sulea et al 2014), and it has continued to gather momentum. More recently, applications of VR have been explored for benefit to patients with advanced cancer.<br><br>Examples:<br>• Pain and anxiety associated with painful medical procedures:<br> ○ VR application for chronic pain: 30 minute intervention, VR headset to enable 'immersion': seems to have promise as a non-opioid treatment and further investigation is warranted (Jones et al 2016)<br>• VR for Management of Pain in Hospitalised Patients – positive results over non VR for reducing pain (Tashjian et al 2017)<br>• Concept of 'Positive Technology': VR designed to increase well-being and to generate strengths and resilience in individuals, organizations, and society:<br> ○ Mood induction in advanced cancer patients: VR to induce positive emotional response for hospital inpatients, preliminary evidence of efficacy in increasing 'positive mood' (Banos et al 2013)<br>• Implementation of VR interventions must take into account the patients' medical state and physical discomfort level, especially with those in the advanced stages of disease (Banos et al 2013).<br><br>**5.2 VR for grief and bereavement**<br>Supporting the needs of family and friends following the death of a patient can be complex, and adapting to the needs of the individual is paramount at this challenging time. Developments in VR have sought to incorporate it into grief and bereavement support, however this has been limited:<br>• VR support group: preliminary efficacy for improving psychosocial outcomes and sleep quality compared to an active control grief education website (Knowles et al 2017).<br><br>**5.3 VR – for education and training needs**<br>VR has the potential to be used to simulate environments for learning opportunities – for example, VR environments |

| | |
|---|---|
|  | have been used to augment surgical training and enable experience of a clinical situation without the need to be physically present.<br><br>Examples:<br>• Potential for VR in communication skills training for clinicians caring for patients with advanced cancer:<br> ○ VR simulation scenarios to meet the learning needs of nurses and health-care professionals caring for men with prostate cancer (Moule et al 2015).<br>• Patient education:<br> ○ mHealth Tool for Lung Cancer (mHealth TLC): A virtual world health game for lung cancer patients – Health games have the potential to improve patient–clinician communication, and mHealth TLC specifically may decrease lung cancer stigma and improve patient-clinician communication (Brown-Johnson et al 2015). |
| **Artificial Intelligence (AI), Machine Learning (ML)** | **Examples/development** |
| | AI and ML have been important areas for research in computing, and it is one of the most dynamic and potential filled areas of technological development (FIT 2018). Developments in AI and ML could revolutionise our relationship (interactions, uses and experience) with technology, including advances in how it integrates into our lives, by creating:<br>• Systems capable of general decision-making and automation outside of narrow specialties.<br>• Automation of human processes.<br><br>Journal – Nature Machine Intelligence:<br>2019 will see the launch of a dedicated journal for Research in AI, machine learning and robotics. Research and development in this area has seen fast paced advances over the last couple of decades. This research has illustrated how artificial intelligence will reinvigorate various technologies, transform society, address longstanding research questions, and how it poses short- and long-term risks for humanity. This journal has been created in recognition of the importance and significance that this area of technology will have on our world.<br><br>**6.1 AI and ML to improve outcomes for individuals: Natural Language Processing (NLP) and systems-based approaches for 'prediction' and 'screening':**<br>Electronic Health Records (EHRs) contain rich information, such as free text entries in patient records as well as clinical assessment and monitoring information, about the physical and psychosocial condition of patients. This information, if brought together and interrogated meaningfully, could provide opportunities for more 'personalised medicine' and improve the way that care is provided and delivered to an individual patient. In recent years there has been a drive to |

harness this ‘tacit’ information, and through the use of MI and ML, interpret and analyse this information.
Natural language processing (NLP) is the ability of a computer program to understand written and spoken language. NLP makes it possible for an artificial intelligence (AI) program to receive conversational input, break syntax down to comprehend meaning, determine appropriate action and respond in a colloquial manner.
Examples:

- ‘Bag-of-Words’ model, utilising free text from EHRs to investigate the potential for ‘feature selection’ strategies (Soguero-Ruiz 2016). EHR’s contain large amounts of longitudinal data that are valuable for biomedical informatics research. ML, such as NLP, offers a promising alternative to manual analysis of heterogeneous temporal data in EHR – “subsequence-based method using a symbolic representation of time series” (Zhao et al 2017).
- AI technology solutions to increase daily physical activity in cancer survivors – Daily motivation from a fully automated, data-driven algorithmic text message via mobile phone (Coachtext); and Voice Assist intervention; in-home on demand autonomous Intelligent Agent using data driven Interactive Digital Voice Assist on the Amazon Alexa/Echo (MyCoach) (RCT – registered 2017, Hassoon et al 2018).
- Prognostication of colorectal cancer patients: Systems-based biomarker discovery approaches – data driven approaches utilising electronically generated health data and machine learning (Vafaee 2018).

**6.2 Role of ‘Big Data’ and AI/ML for Population Health Management – population level data**
Development of care management systems that remain separate from other data sources and patient planning tools is short sighted, precluding future developments.

Application programming interfaces (APIs): This technology enables one software programme to access the services of another, making it possible to track patient data across different venues and enhance care management.

**6.3 AI/ML – automation of human processes: ethical and moral issues**
AI and automation of human processes – replacing or augmenting human interaction. Developments in this area could also take away the need for human interaction, with some fearing that ML/AI has the potential to replace certain job roles with a computer programme. For example the medical transcriptionists, medical records and health information technicians and medical secretaries are the most likely jobs to be computerised in the future (Benedikt Frey and Osbourne 2013), which has huge implications for the employment sector. While commentators suggest that ML and AI technologies cannot replace empathy of a human being, or the non-linear creativity and problem solving abilities of the human brain (The Medical Futurist, 2018), the moral ethics of automating current human processes should be central to any future debates or developments.

| | |
|---|---|
|  | Examples of ML/AI:<br>• Machine Learning to supplement human skills: for example ML for use in medical imaging analytics means a programme could potentially perform the role of human radiologists. Additional benefits include enhanced and timelier scan review, potentially leading to earlier and more accurate diagnoses (Erikson 2017).<br>• Babylon AI Healthcare - Babylon Triage and Diagnostic System (Salman et al 2018):<br> ○ AI powered triage and diagnostic system<br> ○ Illustrates clinical accuracy and comprehensive advice<br> ○ Non human interaction<br> ○ Bias – vignettes cannot be directly interpreted to 'real-world' presentations – based on MRCP clinical exams which prioritise uncommon presentations<br>• Collaboration between ML and medical communities required (Ting et al 2018):<br> ○ Facilitate the development and validation of deep learning techniques<br> ○ Strategy to inform how these technologies can be used to benefit patient care |
| **Robotics** | **Examples/development** |
| | **Robotics**<br>EU 7th Framework funded over 100 collaborative projects on advanced research into robots: developments in understanding the world around them. (https://ec.europa.eu/digital-single-market/en/programme-and-projects/project-factsheets-robotics):<br><br>The European Commission held the 'European Robotics Forum 2018 – Human-Robot-Collaboration in Industry and Services' – March 2018 – presentation of the 7 most successful 7th framework projects in robotics, two examples were related to healthcare: HOBBIT the mutual care robot; RAPP - Empowering robotics for social inclusion.<br>**7.1 Robotics – for assistance**<br>Assistive robotic technology has been expanding – designed to provide assistive support rather than social interaction, for tasks for example: reminders for medication, facilitating skype calls, regulation of room temperature and light levels.<br>• EU 7FP - HOBBIT the mutual care robot – development of an assistive robot that helps seniors and old people at home.<br>• 'Stevie' – an assistive robot for the elderly developed by professor Conor McGinn at Trinity College Dublin<br>• 'Robbie' – assistive robot, developed to be able to recognise 90 common objects as well as human actions and emotions, led by Dr Ardhendu Behera at Edge Hill University. |

**7.2 Robotics – For companionship/social inclusion**
A recent scoping review into the use of 'socially assisted robot' (SAR) technology in the elderly identified that development in this area has the potential to reform delivery of care – studies included showed SAR could improve general mood and well-being, aspects of cognition, and also improve sociability and overcoming the feeling of loneliness and social isolation (Abdi et al 2018).

- RAPP - Empowering robotics for social inclusion - an open source software platform to support the creation and delivery of robotic applications; to increase the versatility and utility of service and assistive robots: applications to enable robots to understand and respond to the intentions and needs of people at risk of exclusion, and especially the elderly.

**7.3 Robotics – optimisation of surgery**
It has been predicted that robotic surgery technology will develop rapidly over the next 10 years. Recent leaps in robotic technology for surgery has made them more versatile, compact and cost-effective. This could lead to the use of robotic surgery more locally, without the need for large hospitals with dedicated robotic suites. This development has the potential to significantly increase the number of surgeries that can be carried out by laparoscopic (keyhole) techniques. This will potentially benefit patients as smaller incisions can reduce the pain of surgery and chance of infection and even contribute to faster recovery times.

Examples include:

- Da Vinci – operates in prostate, bladder and gynaecological surgery, but expanding – however, despite exponential worldwide growth, criticism over industry sponsored studies has been voiced highlighting a lack of clear evidence over other techniques (Criss et al 2018). Key patents currently expired, leaving room for expansion and development by other companies.
- Versius robot surgery system (CMR Surgical) – expected to receive a European health and safety approval mark within the next few months. Versius is smaller, and CMR believes it will be more flexible and versatile than existing robots, allowing it to perform a wider range of operations.
- Google partnered with Johnson & Johnson on medical equipment manufacturer Verb Surgical. The company aims to have its robots connected to the internet so they can learn from each other. Aim to launch 2020.
- Medtronic – another company with plans to launch surgical robots.

**7.4 Robotics – support for education**
Robots could potentially support palliative care education for clinicians and the public. There are some examples of how robot technology has been incorporated into health care education, such as surgery, care of the elderly and social and

| | |
|---|---|
| | behavioural issues, however less so for palliative care education.<br><br>Examples of robotic technology in education include:<br>• There has been evidence for the use of robots to support patient education to reduce reliance on medical professionals (Ishiguro and Majima 2016)<br>• Robots have been used to educate children about the needs of frail elderly people (Masuda et al 2018, Murashima et al 2018)<br>• Social robotics for childhood development, particularly for language development and social behaviours for children with autism (Pennisi et al 2016)<br>• Robotics have been used in combination with virtual reality to develop immersive environments to support surgical training (Bric et al 2016)<br>• **End of life care education:** Robot technology to support the delivery of high fidelity patient simulation (HPS). HPS involves the computerized manikins that simulate real-life scenarios that are safe for teaching and experimentation (Shaw and Abbott 2017)<br><br>Opportunities for robotic technology in palliative care education are great and potentially wide reaching. For example optimising and facilitating social engagement through the use of 'social robotics' to raise awareness about palliative care topics for children, patients, family-caregivers, healthcare professionals and students. Robots also have the potential to enhance the delivery of HPS to improve the education experience and promote 'realism' as they could be programmed to convey emotion, move and respond (autonomously) to the learner. |
| **The Smart Home** | **Examples/development** |
| | A Smart Home: a residence equipped with computing and information technology which anticipates and responds to the needs of the occupants, through the management of technology within the home and connections to the world beyond (Solaimani et al 2015).<br><br>Objective: 'informational' home, where existing and new information services are interactively connected to the outside world, rather than the mere 'automation' of home appliances.<br><br>**8.1 Smart Home Sensors – detect changes in health condition/physical function**<br>Potential to use Smart Home data as 'alerts' or 'flagging' through 'ambient sensors' for automated activity. Potential to quantify and explain changes that are detected in daily activity data: activity recognition using deep neural networks.<br><br>Studies have reported positive outcomes, using remote assessment technologies such as web/smart phone-based self- |

reports and wearable sensors, however the cancer research community is still lacking far behind. Thorough investigation of more advanced technologies in cancer care is warranted (Fallahzadeh et al 2018).
Examples:

- Behaviour Change Detection (BCD):
  - Older adults living in smart homes who experienced major health events, including cancer treatment, insomnia, and a fall (Sprint et al 2016 conference presentation, Sprint et al 2017)
  - On-body inertial sensors – detection of changes in physical and cognitive health (Bulling et al 2014)
  - SHARON simulator – synthetic dataset predicting decline in patients with Alzheimer's Disease (Masciadri et al 2017)
  - Future research focus – studies to adapt BCD to analyse smart phone and wearable data, as well as data collected in smart homes (Sprint et al 2017).
- Wireless Body Area technology:
  - Ambient Assisted Living for All project: novel ambient assisted-living environment learn human behaviour and, over time, perform desired actions without human intervention. Such systems have the potential to supplement care for the elderly or physically impaired – allowing remote monitoring by informal or formal caretakers (Costa et al 2014).
  - Ambient Assisted Living (AAL) systems: identify and predict situations that may endanger users in their living environment developed with the principles of reactive and proactive behaviour validated with a case study and supported by a probabilistic ontology (Machado et al 2017).
- Gator Tech Smart House (GTSH) – Professor Sumi Helal (Helal 2009) - real-world research facility ('smart space') to test 'pervasive technology' designed for older and disabled people. Project goal: "create assistive environments such as homes that can sense themselves and their residents and enact mappings between the physical world and remote monitoring and intervention services" (Helal 2005).

**8.2 Smart Home Sensors – Alert systems and monitoring – home security and controls**

The use of smart home sensors for home security has been an area of rapid growth. Feeling insecure in our homes, can have a detrimental impact on our health and wellbeing. Physical decline from long term conditions or serious illness can also impact on how we feel in our homes and how we 'live' within them.
Innovations have focussed on every day use, however the potential exists for applications to healthcare. For example, providing a mechanism for an individual to monitor and control their living environment may benefit people who have problems with mobility.

Examples:

| | |
|---|---|
| | • Smart doorbells – alert to who is at the door, including keyless locking and unlocking – companies: Everest, Safe, Nuki, Ring, Google Nest<br>• Voice recognition (Alexa/Echo)<br><br>**8.3 Smart Cities – Built Environment and Big Data**<br>Cities have the potential to pull information from many sources in order to understand more about its residents and their needs. For example data from mobile device sensors and ambient sensors, data can also be tapped from citywide sites such as power grid status, transportation grid status, vehicular networks, NHS and healthcare service data etc. Strategies for city planning and the built environment should include the optimisation and use of 'smart' technology to generate data that can be used to improve the health of the resident population (Cook et al 2018).<br><br>Population Health Management: 'big data' from 'smart cities' has the potential to shape the health of communities and to rethink how health and care services can be delivered.<br>Examples:<br>○ NHS – 10 Healthy New Towns (https://www.england.nhs.uk/ourwork/innovation/healthy-new-towns/)<br>○ Future investment is required for research into using Smart City Technology to make healthcare smarter (Cook et al 2018) |
| **Biotechnologies/Genome Editing** | **Examples/development** |
| | "Truly one-of-a-kind"– Technology exists to enable a comprehensive "omic" assessment of an individual at a biological level (e.g. DNA sequencing). This understanding presents a remarkable and unprecedented opportunity to improve medical treatment and develop preventive strategies to preserve health (Topol 2015).<br><br>Most uses of genome editing have so far been in scientific research, for example to investigate models of human disease. Given that genome editing has the potential to alter any DNA sequence, whether in a bacterium, plant, animal or human being, it has an almost limitless range of possible applications in living things.<br><br>**9.1 Genome profiling and Personalised/Individualised Medicine**<br>Understanding an individual's genetic makeup offers the potential to tailor treatment and medications to that individual. This could enable professionals to predict which medical treatments would be safe and effective for an individual |

| | |
|---|---|
| | patient, and which ones would not be (Mathur and Sutton 2017).<br><br>Potential benefits to palliative care include, for example, more targeted use of opiods tailored to the patient based on genetic profile leading to more effective pain control and reduction of problems associated with opioid use such as opioid toxicity and unwanted side effects. Research however is very limited, especially in palliative care.<br><br>**9.2 Genetic editing and biomarker technology for earlier disease detection and possible disease prevention**<br>Genome editing techniques are now widely used in research across many areas of human health and offer prospects for treating disease, avoiding genetic diseases, and human enhancement. Genetic variations may not always directly cause disease, but may be associated with an increased risk of developing a certain disease or, conversely, have a protective effect against a certain disease (Nuffield Council on Bioethics).<br><br>Despite consensus guidelines outlining that genome editing is justified for scientific purposes (fundamental biology), this raises ethical issues due to the possibility that scientists could make permanent modifications to the human germ line (Editorial, Nature 2017). |
