## Supplementary material for "Technology in Palliative Care (TIP): the identification of digital priorities for palliative care research using a modified Delphi method": Summary of networks used to invite palliative care professionals to participate

**Appendix: Summary of the networks used to invite palliative care professionals to participate**

| Group(s) contacted | Method | Approximate reach (n) |
|---|---|---|
| International Collaborative for Best Care of the Dying Person (a multidisciplinary, international palliative care research group; https://bestcareforthedying.org) | Individuals contacted by email via administrator | 79 |
| CHAIN (Contact, Help, Advice and Information Network; https://www.networks.nhs.uk/nhs-networks/chain) UK National Health Service (NHS) technology interest email list. | Individuals contacted by email via administrator | 8326 |
| Nine Marie Curie Hospices UK | Individuals contacted by email via local administrators and research leads. | 450 |
| NHS England email distribution lists of the following: Allied Health Professionals (National); Healthcare Science Practitioners | Individuals contacted by email via administrator | 1000 |

| | | |
|---|---|---|
| (National); Psychological professions (North); Cancer alliances (North); Genomics network (National); Informatics and bioinformatics (North West). | | |
| Social media | Four tweets on Twitter were sent from ACN's Twitter profile. | 12058 |
| Technology in Palliative Care Special Interest Group (https://amaranwosu.com/pads) | Targeted email to professionals from a palliative care technology event who had previously provided consent to receive information about future studies | 57 |
