## Supplementary material for "Technology in Palliative Care (TIP): the identification of digital priorities for palliative care research using a modified Delphi method": Voting outcomes for consensus meeting

**Appendix: Voting outcomes for consensus meeting**

| High Agreement | | Group 1 | Group 2 | Deciding vote outcome (n=11) |
|---|---|---|---|---|
| 1 | Development of Electronic Health Records (EHR) and systems-based approaches to collection and utilisation of Big Data | In | In | NA |
| 2 | Governance in the use of 'Big Data' – regulation and responsibility | In | In | Included combined with item 3 |
| 3 | Cybersecurity and ransomware – Packaging, capturing and transferring data and keeping it safe | In | Out | Included (combined) 11/11<br><br>Combined with item 2 |
| 4 | Ethical Challenges in Big Data Health Research: implications for informed consent and participation | In | Out | 11/11 |

| | | | | |
|---|---|---|---|---|
| 5 | Use of mobile devices to gather patient reported symptom outcomes (PRO's)/Patient Performance Status | In | In | NA |
| 6 | App design: Clinical input – Safety, efficacy, accuracy and assessment of risk | In | In | NA |
| 7 | Telehealth/eHealth to support patients and their families in their own homes | In | In | Combined with item 8 |
| 8 | Telehealth/eHealth for electronic patient-reported outcomes (ePRO) | In | Out | Included 11/11<br><br>combined with item 7 |
| 9 | VR: for education and training needs | Out | Out | |
| **Moderate Agreement** | | **Group 1** | **Group 2** | **Vote / Outcome** |
| **10** | Patient-Generated Health Data (PGHD) – what to collect and how: harnessing and using 'physical function' data via sensor-based technologies (edited) | Out | In | Included 11/11 |
| **11** | Smart Home Sensors – detect changes in health condition/physical function | Out | In | Included 6/11 |

| Low Agreement | | Group 1 | Group 2 | Vote / Outcome |
|---|---|---|---|---|
| **12** | Smart Threads – early development experimental stages but with positive results as a potential diagnostic or monitoring technology, or as a mechanism for displaying information | Out | Out | NA |
| **13** | Physical Activity Change Detection (PACD) | Out | In | Included (combined) 10/11 combined with item 15 |
| **14** | Reducing burdensome interventions for patients: optimising monitoring and recording of physiological signals | Out | Out | NA |
| **15** | Wearable Health Trackers | Out | In | Included (combined) 9/11 combined with item 13 |
| **16** | Wearable fabrics for the detection of stimuli (physical functioning e.g. temperature, motion, strain, activity monitoring) | Out | Out | NA |

| | | | | |
|---|---|---|---|---|
| **17** | Body area network (BAN) technology | Out | Out | NA |
| **18** | Wearable technology – privacy and ethical considerations | Out | Out | NA |
| **19** | Portable Hospital-level screening/diagnostics in the home | Out | Out | NA |
| **20** | VR: Distraction therapy to ameliorate symptoms – pain, distress, anxiety | In | In | NA |
| **21** | VR: grief and bereavement following the death of a patient | Out | Out | NA |
| **22** | AI and ML to improve outcomes for individuals: Natural Language Processing (NLP) and systems-based approaches for ‘prediction’ and ‘screening | In | In | NA |
| **23** | Role of ‘Big Data’ and AI/ML for Population Health Management – population level data | In | In | NA |
| **24** | AI/ML – automation of human processes: ethical and moral issues | In | In | NA |
| **25** | Robotics – for assistance and daily living | Out | Out | NA |
| **26** | Robotics – for companionship/social inclusion | Out | Out | NA |
| **27** | Robotics – optimisation of surgery | Out | Out | NA |

| | | | | |
|---|---|---|---|---|
| **28** | Robotics – Education (including simulation) | Out | Out | NA |
| **29** | Smart Home Sensors – Alert systems and monitoring – home security and controls | Out | Out | NA |
| **30** | Smart Cities – Built Environment and Big Data | Out | Out | NA |
| **31** | Genome profiling and Personalised Medicine | In | In | NA |
| **32** | Genetic editing and biomarker technology for earlier disease detection and possible disease prevention | In<br>Focus on biomarkers rather than genetic editing. | Out | Included 11/11 |
