## Supplementary material for "Technology in Palliative Care (TIP): the identification of digital priorities for palliative care research using a modified Delphi method": Delphi questionnaire - Google form

### Technology in Palliative care (TIP) Study

Questions Responses 167

Section 1 of 4

#### Technology in Palliative Care (TIP) Study: Priority Setting to Improve the Care of Patients with Advanced Cancer

Form description

Email address *

Valid email address

This form is collecting email addresses. Change settings

##### Consent To Participate - Round 2 Delphi Questionnaire

Please read the participant information sheet before confirming your consent to participate - https://drive.google.com/open?id=1xJNuuHneXPzBQPCUX28w9lzq39aD3uUT

##### Title

Please read each statement and then tick to confirm your agreement.

1. I confirm that I have read and have understood the information sheet dated 22nd *

Tick to confirm

2. I understand that my participation is voluntary and that I am free to withdraw at any time without giving any reason, without my rights being affected. In addition, should I not wish to answer any particular question or questions, I am free to decline. *

Tick to confirm

3. I understand that, under the Data Protection Act 1998, I can at any time ask for access to the information I provide and I can also request the destruction of that information if I *

Tick to confirm

4. I agree for the data I provide to be archived at the University of Liverpool. I understand that other authorised researchers will have access to this data only if they agree to preserve the confidentiality of the information as requested in this form. *

Tick to confirm

5. The information you have submitted will be published as a report; please indicate whether you would like to receive a copy.

Tick to confirm

6. I understand that the researcher will contact me to ask if I would be prepared to participate in further elements of this study. *

Tick to confirm

7. I agree to take part in the above study *

After section 1 Go to section 2 (Technology in Pall...mographic Details)

Section 2 of 4

### Technology in Palliative Care (TIP) Study: Demographic Details

Description (optional)

Country of residence *

Short answer text

Age (please enter numerically) *

Short answer text

Gender *

- Female
- Male
- Other

Occupation (including field of expertise) *

Short answer text

After section 2

Continue to next section

Section 3 of 4

### Technology in Palliative Care (TIP) Study: Priority Setting to Improve the Care of Patients with Advanced Cancer: Round 2 Delphi Questionnaire

Your answers to the questionnaire in both rounds of this Delphi study will generate 'priority areas' to focus research and development into the use of digital technology, with the aim of improving the care experience of patients with advanced cancer

Before completing the questionnaire please review the Round 1 Delphi questionnaire results summary document, as well as the 'Scoping Review' document which provides further information around each theme area. The complete Scoping Review can be viewed and/or downloaded by clicking on the link below. Additionally, links to the specific sections of the scoping review are provided for each theme area of the questionnaire.

SCOPING REVIEW: https://drive.google.com/open?id=1ioHUZc89p0pXDEQyedjEdCdAPenlftMG

#### Each category presented below represents a potential priority area, to focus future research and development, to improve the care of patients with advanced cancer. Please indicate the level or priority you think should be given to each one, on a scale of 1-5.

To help you complete this questionnaire, a link to the specific section of the scoping review can be found under each section heading

#### Big Data: 1.1 - 1.5

Big Data provides an opportunity to gather large amounts of data on the health of the population, but there are also implications for data transfer and how this data is used. Please indicate the level of priority that should given to each of the following:

#### Title

Section 1: https://drive.google.com/open?id=1fHTFjp-5YZnLzQSrfIZxsPSX9815hRNt

1.1 Development of Electronic Health Records (EHR) and systems based approaches to collection and utilisation of Big Data *

Low Priority ○ ○ ○ ○ ○ High Priority

1.2 Patient-Generated Health Data (PGHD) and Big Data – what to collect and how: harnessing and using 'physical function' data via sensor-based technologies *

| | 1 | 2 | 3 | 4 | 5 | |
|---|---|---|---|---|---|---|
| Low Priority | ○ | ○ | ○ | ○ | ○ | High Priority |

1.3 Governance in the use of 'Big Data' – regulation and responsibility *

| | 1 | 2 | 3 | 4 | 5 | |
|---|---|---|---|---|---|---|
| Low Priority | ○ | ○ | ○ | ○ | ○ | High Priority |

1.4 Cybersecurity and ransomware – Packaging, capturing and transferring data and keeping it safe *

| | 1 | 2 | 3 | 4 | 5 | |
|---|---|---|---|---|---|---|
| Low Priority | ○ | ○ | ○ | ○ | ○ | High Priority |

1.5 Ethical Challenges in Big Data Health Research: implications for informed consent and participation *

| | 1 | 2 | 3 | 4 | 5 | |
|---|---|---|---|---|---|---|
| Low Priority | ○ | ○ | ○ | ○ | ○ | High Priority |

Wearable's and Sensor-Based Technologies (on the body): Assessment of physical function and [illegible]

The Future Institute Today tracker shows 538 wearable devices designed to using sensor based technologies. Please consider each of the following sub-categories and indicate the level of priority that should given to each of the

Title

Section 2: https://drive.google.com/open?id=1_vPVCli-QHMNhUyUyPh2EMMfNe4Nnm1M

2.1 Smart Threads – early development experimental stages but with positive results as a potential diagnostic or monitoring technology, or as a mechanism for displaying *

| | 1 | 2 | 3 | 4 | 5 | |
|---|---|---|---|---|---|---|
| Low Priority | ○ | ○ | ○ | ○ | ○ | High Priority |

2.2 Physical Activity Change Detection (PACD) *

| | 1 | 2 | 3 | 4 | 5 | |
|---|---|---|---|---|---|---|
| Low Priority | ○ | ○ | ○ | ○ | ○ | High Priority |

2.3 Reducing burdensome interventions for patients: optimising monitoring and recording of physiological signals *

| | 1 | 2 | 3 | 4 | 5 | |
|---|---|---|---|---|---|---|
| Low Priority | ○ | ○ | ○ | ○ | ○ | High Priority |

2.4 Wearable Health *

| | 1 | 2 | 3 | 4 | 5 | |
|---|---|---|---|---|---|---|
| Low Priority | ○ | ○ | ○ | ○ | ○ | High Priority |

motion, strain, activity monitoring)

| | 1 | 2 | 3 | 4 | 5 | |
|---|---|---|---|---|---|---|
| Low Priority | ○ | ○ | ○ | ○ | ○ | High Priority |

2.6 Body area network (BAN) technology *

| | 1 | 2 | 3 | 4 | 5 | |
|---|---|---|---|---|---|---|
| Low Priority | ○ | ○ | ○ | ○ | ○ | High Priority |

2.7 Wearable technology – privacy and ethical considerations *

| | 1 | 2 | 3 | 4 | 5 | |
|---|---|---|---|---|---|---|
| Low Priority | ○ | ○ | ○ | ○ | ○ | High Priority |

#### Mobile Devices: 3.1 - 3.3

Mobile devices have become ubiquitous and integral to the way that we conduct our lives. From laptops, hand held devices, mobile diagnostic and clinical devices to name a few, these advances in technology have altered the way in which healthcare data can be collected as well as the way clinical procedures and patient monitoring can be carried out. Please consider the following ways mobile technology could support care and indicate the level of priority that should given to each of the following:

#### Title

Section 3: https://drive.google.com/open?id=1qLUC9ofylxYqIoImZQcbElVMWDwibttD

3.1 Use of mobile devices to gather patient reported symptom outcomes (PRO's)/Patient Performance Status *

| | 1 | 2 | 3 | 4 | 5 | |
|---|---|---|---|---|---|---|
| Low Priority | ○ | ○ | ○ | ○ | ○ | High Priority |

3.2 App design: Clinical input – Safety, efficacy, accuracy and assessment of risk *

| | 1 | 2 | 3 | 4 | 5 | |
|---|---|---|---|---|---|---|
| Low Priority | ○ | ○ | ○ | ○ | ○ | High Priority |

3.3 Portable Hospital-level screening/diagnostics in the home *

| | 1 | 2 | 3 | 4 | 5 | |
|---|---|---|---|---|---|---|
| Low Priority | ○ | ○ | ○ | ○ | ○ | High Priority |

#### Telehealth/eHealth: 4.1 - 4.2

Telehealth and eHealth have the potential to impact on how patients and health professionals interact, by utilising online and remote communications. Please indicate the level of priority that should given to each of the following:

#### Title

Section 4: https://drive.google.com/open?id=1ThZzfcurR_S7U1JMhaxSuYsHfmBIDn0i

4.1 Telehealth/eHealth to support patients and their families in their own homes *

| | 1 | 2 | 3 | 4 | 5 | |
|---|---|---|---|---|---|---|
| Low Priority | ○ | ○ | ○ | ○ | ○ | High Priority |

4.2 Telehealth/eHealth for electronic patient-reported outcomes (ePRO) *

| | 1 | 2 | 3 | 4 | 5 | |
|---|---|---|---|---|---|---|
| Low Priority | ○ | ○ | ○ | ○ | ○ | High Priority |

VR has a wide range of applications that could benefit patients with advanced cancer, from therapeutic interventions for symptom control to supporting the education of health professionals. Please indicate the level of priority that should given to each of the following:

#### Title

Section 5: https://drive.google.com/open?id=1fCkwBbnSql4-7VOsgV4c805yFvmxMp8x

##### 5.1 VR: Distraction therapy to ameliorate symptoms – pain, distress, anxiety *

| | 1 | 2 | 3 | 4 | 5 | |
|---|---|---|---|---|---|---|
| Low Priority | ○ | ○ | ○ | ○ | ○ | High Priority |

##### 5.2 VR: grief and bereavement following the death of a patient *

| | 1 | 2 | 3 | 4 | 5 | |
|---|---|---|---|---|---|---|
| Low Priority | ○ | ○ | ○ | ○ | ○ | High Priority |

##### 5.3 VR: for education and training needs *

| | 1 | 2 | 3 | 4 | 5 | |
|---|---|---|---|---|---|---|
| Low Priority | ○ | ○ | ○ | ○ | ○ | High Priority |

#### Artificial Intelligence (AI), Machine Learning (ML): 6.1 - 6.3

AI and ML have the potential to change the way we deliver care for patients with advanced cancer, offering unique opportunities for population health through innovations in screening and health monitoring. Please indicate the level of priority that should given to each of the following:

#### Title

Section 6: https://drive.google.com/open?id=1pYcQUBBWoA2LNWRk2t484mqaqazHc7Kj

systems-based approaches for prediction and 'screening'

| | 1 | 2 | 3 | 4 | 5 | |
|---|---|---|---|---|---|---|
| Low Priority | ○ | ○ | ○ | ○ | ○ | High Priority |

6.2 Role of Big Data and AI/ML for Population Health Management – population level data *

| | 1 | 2 | 3 | 4 | 5 | |
|---|---|---|---|---|---|---|
| Low Priority | ○ | ○ | ○ | ○ | ○ | High Priority |

6.3 AI/ML – automation of human processes: ethical and moral issues *

| | 1 | 2 | 3 | 4 | 5 | |
|---|---|---|---|---|---|---|
| Low Priority | ○ | ○ | ○ | ○ | ○ | High Priority |

#### Robotics: 7.1 - 7.4

The development of robotic technology is set to increase dramatically over the next 10 years, with innovations in diverse care settings such as care homes for dementia patients and keyhole surgery in hospital theatres. Please indicate the level of priority that should given to each of the following:

#### Title

Section 7: https://drive.google.com/open?id=1LsDSCc26l2DM8asni6ovDF0q7KmkjuSd

7.1 Robotics – for assistance and daily living *

| | 1 | 2 | 3 | 4 | 5 | |
|---|---|---|---|---|---|---|
| Low Priority | ○ | ○ | ○ | ○ | ○ | High Priority |

| | 1 | 2 | 3 | 4 | 5 | |
|---|---|---|---|---|---|---|
| Low Priority | ○ | ○ | ○ | ○ | ○ | High Priority |

7.3 Robotics – optimisation of surgery *

| | 1 | 2 | 3 | 4 | 5 | |
|---|---|---|---|---|---|---|
| Low Priority | ○ | ○ | ○ | ○ | ○ | High Priority |

7.4 Robotics – Education *

| | 1 | 2 | 3 | 4 | 5 | |
|---|---|---|---|---|---|---|
| Low Priority | ○ | ○ | ○ | ○ | ○ | High Priority |

#### The Smart Home: 8.1 - 8.3

The integration of sensor based technology into the everyday environment offers wide reaching opportunities for healthcare monitoring and provision. Please indicate the level of priority that should given to each of the following:

#### Title

Section 8: https://drive.google.com/open?id=1DnDxIXWpPHVPCdFFn3nRk1PBtGGVEdhz

8.1 Smart Home Sensors – detect changes in health condition/physical *

| | 1 | 2 | 3 | 4 | 5 | |
|---|---|---|---|---|---|---|
| Low Priority | ○ | ○ | ○ | ○ | ○ | High Priority |

8.2 Smart Home Sensors – Alert systems and monitoring – home security and controls *

| | 1 | 2 | 3 | 4 | 5 | |
|---|---|---|---|---|---|---|
| Low Priority | ◯ | ◯ | ◯ | ◯ | ◯ | High Priority |

##### 8.3 Smart Cities – Built Environment and Big Data *

| | 1 | 2 | 3 | 4 | 5 | |
|---|---|---|---|---|---|---|
| Low Priority | ◯ | ◯ | ◯ | ◯ | ◯ | High Priority |

#### Biotechnologies/Genome Editing/Profiling: 9.1 - 9.2

Biotechnologies and advances in genome profiling and editing has the potential to revolutionise the healthcare landscape, from targeted medicine's based on individual gene profiles, to modifications of the human genome. Please indicate the level of priority that should given to each of the following:

#### Title

Section 9: https://drive.google.com/open?id=1VQKaHaKpUFZ8s-3-4cMgnNgiTMGKGJRz

##### 9.1 Genome profiling and Personalised Medicine *

| | 1 | 2 | 3 | 4 | 5 | |
|---|---|---|---|---|---|---|
| Low Priority | ◯ | ◯ | ◯ | ◯ | ◯ | High Priority |

##### 9.2 Genetic editing and biomarker technology for earlier disease detection and possible disease prevention *

| | 1 | 2 | 3 | 4 | 5 | |
|---|---|---|---|---|---|---|
| Low Priority | ◯ | ◯ | ◯ | ◯ | ◯ | High Priority |

After section 3 Continue to next section

### Development of an International Collaboration for Technology in Palliative Care (InC-TIP)

Description (optional)

I am happy to be invited to be part of the International Collaboration. Please indicate below: *

- Yes
- No
