## Supplementary material for "Technology in Palliative Care (TIP): the identification of digital priorities for palliative care research using a modified Delphi method": Examples of technologies used in palliative care during the COVID19 pandemic

**Appendix: Examples of use of technologies in palliative care during the COVID19 pandemic**

| Example | Author | Application |
|---|---|---|
| **Support patient care and assessment with telehealth** | Calton et al [1] | Developed recommendations for palliative care clinicians to deliver telemedicine in clinical practice. |
| | Grewal et al[2] | Telehealth used to remotely monitor chemotherapy response in a 75-year-old woman. |
| | Ritchey et al[3] | Telehealth used to facilitate remote assessment and to support family discussion for a patient dying from COVID-19. |
| | Bettini et al [4] | Facilitation of goals-of-care discussions and at end of life for an infant in the intensive care unit. |
| | Chávarri-Guerra[5] | Telehealth used to deliver remote interventions to oncology patients (psychological care, pain and symptom management, nutrition assessment, physical therapy, end of life care, geriatric assessment, advance directive completion and psychiatric care. |

| | |
|---|---|
| Mackey et al[6] | Telehealth (InTouch platform) was used for remote assessment of a patient with COVID-19 infection. |
| Harris et al[7] | Telehealth was used to provide remote assessments for residents in long term care facilities. |
| Flores et al[8] | Used telemedicine to support remote palliative care assessments in an hospital emergency department. |
| Lu et al[9] | Used a remote and wireless programming system to perform video-based real-time programming and pain management for patients with a implanted spinal cord stimulation. |
| Sansom-Daly [10] | Telehealth-delivered outpatient clinical psychology practice with adolescent and young adults with cancer. |
| Samara et al[11] | Telehealth was used by nurse practitioners to provide palliative care for long term care residents. |
| Chua et al[12] | Identified key elements of ‘webside manner’ that are helpful when conducting serious illness conversations by virtual visit. |

| | | |
|---|---|---|
| | Lally K et al[13] | Describe how the palliative care clinic at cancer centre was converted into a predominantly telemedicine model. |
| **Communication between healthcare professionals** | Crosby et al[14] | Presented the views of hospital-based specialist palliative care staff who were using video communication technology during the pandemic. |
| **Virtual communities of practice** | Mill et al[15] | Described the emergence of online resources and virtual communities of practice to support palliative care delivery during the pandemic. |
| | Abel and Taubert[16] | Provided examples of how technologies have been used to maintain compassion in communities. |
| **Analysis online information to determine public sentiment** | Selman et al[17] | Explored the views and experiences of Twitter social media users who reported that a relative, friend or acquaintance died of COVID-19 without a family member/friend present. |
| | Selman et al[18] | Document analysis of UK online newspaper articles published during 2 week-long periods in March–April 2020, |
| **Virtual reality** | Niki et al[19] | Provided guidance of how VR can be used to conduct virtual travel and family connection in palliative care during the pandemic, |

| | | |
|---|---|---|
| | Wang et al[20] | Highlighted how VR technology can potentially provide psychological care and comfort for patients and their families by enabling remote interaction with dying patients or fulfilling dying wishes. |
| | Niki et al[21] | Highlighted how virtual reality reminiscence reduces anxiety for elderly patients. |
| | Posner et al[22] | Described how virtual reality used to deliver training video for the general public to instruction to use personal protective equipment correctly, when going into palliative care settings to provide care for their family members. |
| **Education and training** | Lal et al[23] | Describe how video communication technology was used to train medical students through observation of outpatient telemedicine and tele-palliative care clinics. |
