## Supplementary material for "Technology in Palliative Care (TIP): the identification of digital priorities for palliative care research using a modified Delphi method": Future Today Institute Trends 225 list

### TABLE OF CONTENTS


02 **Welcome Letter**

08 **Executive Summary**

08 **Key Takeaways**

10 **Guide To The Year Ahead**

12 **Methodology**

15 **How To Use Our Report**

16 **Ten Important Questions**

22 **The Most Important Tech Trends For Your Industry And Organization**

47 **Artificial Intelligence**
51 The AI Cloud
51 Proprietary, Homegrown AI Languages
51 AI Chipsets
52 Cognitive Computing
52 Bots
52 Marketplaces For AI Algorithms
52 More Consolidation in AI
53 Consumer-Grade AI Applications Debut
53 Digital Assistants Become Ubiquitous
53 A Bigger Role For Ambient Interfaces
53 Deep Linking Everywhere
54 Making AI Explain Itself
54 Accountability and Trust
54 Hidden Bias Leads To Big Problems
55 China's AI Boom
56 Real-Time Machine Learning
56 Natural Language Understanding (NLU)
56 Machine Reading Comprehension (MRC)
56 Natural Language Generation (NLG)
56 Generative Algorithms For Voice, Sound and Video
57 Image Completion
57 Predictive Machine Vision
57 Much Faster Deep Learning
57 Reinforcement Learning and Hierarchical RL
57 Continuous Learning
58 Multitask Learning
58 Adversarial Machine Learning

59 **Recognition Technologies**
60 Faceprints
61 Voiceprints
62 Wifi Tracking
63 Gesture Recognition
64 Personality and Character Recognition
65 Ambient Proximity
66 Hidden Bias in Recognition Technologies

67 **Security, Privacy and Data**
69 Compliance Challenges and Unrealistic Budgets
69 DDoS Attacks Will Increase
69 Ransomware As A Service
70 Russia's Gifted Hacker Community Grows
70 New Infrastructure Targets
70 Hacktivism On The Rise
71 Third-Party Verified Identities
71 Targeted Attacks on Digital Assistants
71 Zero-Knowledge Proofs Go Commercial
71 Zero-Day Exploits On The Rise
71 Backdoors
73 Remote Kill Switches
73 AI-Powered Automated Hacking
73 Offensive Government Hacking
73 More Cyber Mission Forces in the Field
73 Strange Computer Glitches Will Keep Happening
74 Proliferation of Darknets, Aided By Cryptocurrencies
74 New Open Source App Vulnerabilities
74 Selfie Security Using Faceprints
74 Bounty Programs
79 The General Data Protection Regulation Takes Effect
79 Right To Eavesdrop/ Be Eavesdropped On
79 Defining What Constitutes Online Harassment
80 Drone Surveillance
80 Personal and One-To-Few Networks
80 Leaking
80 Blocking the Ad Blockers
81 Organizational Doxing
81 Anonymity
81 Authenticity
81 Differential Privacy
82 Anti-Recognition Cammo and Glasses
82 Digital Self-Incrimination
82 SWATting at Trolls
83 Revenge Porn
83 Eye In The Sky
83 Law Enforcement Using Recognition Algorithms To ID Faces
84 Data Retention Policies
84 Encryption Management


© 2018 FUTURE TODAY INSTITUTE

84 Who Owns Your Personal, Biometric Data?
85 Global Data Scientist Shortages

**86 Advanced Robotics**
87 Self-Assembling Robots
88 Robot Compilers
89 Molecular Robotics
90 Collaborative Robotics
91 Ethical Manufacturing
92 Soft Robotics
93 Human-Machine Interfaces
94 Smart Dust
95 Personal Robots and Butlers
97 Robot Abuse
98 3D Printing

**99 Transportation**
100 Flying Taxis
100 Autonomous Underwater Vehicles (AUVs)
100 Drone Delivery
101 Increasing Patents and Calls For New Regulation
102 Drone Lanes
102 Personal Home Drone Surveillance
102 Sense And Avoid Technology
102 Microdrones and Drones Used In Dangerous/Hard-To-Reach Areas
102 Drone Swarms
102 Clandestine, Disappearing Drones
103 Autonomous Vehicle Testing In Cities Around the U.S.
103 Armchair AV Mechanics
103 Assisted Driving Before Full Automation
104 Adaptive Driving Systems
104 Vehicle-to-Vehicle (V2V) Communications
104 Electric Vehicles
104 Cars as Interfaces
104 Solar Highways
105 Autonomous Vehicle Legislation
105 Flying Cars
106 Flights
107 Autonomous Ships

**108 News Media, Book Publishing, Social Networks and the First Amendment**
110 Natural Language Generation for Reading Levels
111 Computational Photography
112 Computational Journalism
113 I-Teams For Algorithms and Data
114 Voice Interfaces For News and Books
115 Proximity News
116 Crowdlearning
117 Digital Frailty
119 Radical Transparency
120 Limited-Edition News Products
121 One-To-Few Publishing
122 Notification Layer (A Tragedy of the Commons)
123 Journalism as a Service (JaaS)
124 Transparency in Metrics
125 Real-Time Fact Checking
126 Offline Is The New Online
127 Audio Search Engines
128 Synthetic Data Sets
129 Connected TVs
129 Decentralizing The Web
129 Streaming Social Video
130 New Video and Audio Story Formats
131 Media Consolidation
132 Tweaks To Social Network Algorithms
133 The First Amendment in a Digital Age

**134 Entertainment Media**
135 Holograms
135 Virtual Reality
136 360-degree Video
136 Augmented Reality
137 Mixed Reality Arcades
138 MMOMRGs

**141 Marketing and Advertising Technologies**
142 VR For Marketing
143 AI For the Creative Process
144 FOBO
145 Retail APIs
146 Digital Associates

**147 Energy**
148 Green Tech
149 Charging Stations
150 Ultra-High-Voltage Direct Current and Macro Grids
151 Better Batteries

**152 Climate And Geoscience**
153 Anthropocene
155 Extreme Weather Events
157 Human Migration Patterns Shift
158 Geoengineering

**159 Agricultural Technologies**
160 Indoor and Outdoor Plant Factories and Microfarms
161 Deep Learning For Farming and Food Recognition
162 Smart Farms
163 Terraforming
164 Cultivated Food and Beverage

**165 Biotechnologies, Genomic Editing and Biointerfaces**
170 Genome Editing
170 Biological DVRs
170 Human DNA-Powered Devices
170 Using Our DNA As Hard Drives
170 Nanobot Nurses
170 Custom-Crafted Microbes

© 2018 FUTURE TODAY INSTITUTE

171 Precision Medicine Just For You
171 Running Out Of Space For Genome Storage
171 Genome Editing Research Clashes With Public Opinion
171 Nootropics and Neuroenhancers
172 Microbiome Extinction
173 Building A Comprehensive Human Cell Atlas
174 Biointerfaces Laminated Onto Our Skin

**175 Health Technologies and Wearables**
177 Patient-Generated Health Data
178 Touch-Sensitive Prosthetics
179 Smart Thread
180 Bioelectronics
181 Wearables
181 Smart Glasses
181 Hearables / Earables
181 Head Mounted Displays
181 Smart Bras
182 Smart Helmets
182 Smart Gloves
182 Tattooables
183 Thinkables
183 Ingestables
183 Smart Fabrics
183 Smartwatches
183 Smart Shoes
183 Wireless Body Area Networks

**184 Smart Homes and the Internet Of Things**
186 Privacy Laws, Net Neutrality and Hackers Threaten the Internet of Things
188 Searching The Internet of Physical Things
189 Intelligent Cameras
190 Our Appliances Will Have Digital Assistants
190 Smart Appliance Screens Are Coming
190 Home Appliances Will Talk To Each Other
190 Wireless Kitchens
191 Smarter Home Security
191 Smart Remotes
191 Smart Mirrors
191 Our Smarthomes Become Weaponized

**192 Workplace and Learning Technologies**
193 Universal Basic Income (UBI)
194 AI in Hiring
196 Productivity Bots
198 Adaptive Learning
199 Nanodegrees
200 Sharing Economy & Lendership

**201 Financial Technologies and Cryptocurrencies**
202 E-Resident and Location-Independent Digital Business Identity Cards
204 Social Payments
205 Cryptocurrencies
207 Blockchain
208 Open Banking
209 Financial Inclusion and Targeting the Underbanked

**210 Smart Cities**
213 Smart City Initiatives
214 Faster Connectivity With 5G
215 City-Level Cyber Security

**217 Government And Technology Policy**
218 Splinternets
219 Election Security
220 Anti-Trust Lawsuits
221 Old Laws Clash With New Technology
223 Digital Caliphate
224 Governments Asking Tech Companies To Help Fight the Spread of Misinformation, Propaganda and Terrorism
225 Overhauling Government Tech Infrastructure

**227 Space**
228 Commercial Space Flight
229 CubeSats
230 Asteroid Mining For Resources
231 Space Exploration

**232 Ten Weak Signals For 2019**

**235 About the Authors**

**236 About The Future Today Institute**

**237 Disclaimer**

**238 The Signals Are Talking**

**239 Companies, Organizations, Universities and Government Agencies Mentioned In Our 2018 Trends Report.**

**247 Contact Information**

© 2018 FUTURE TODAY INSTITUTE 
