## Supplementary material for "Technology in Palliative Care (TIP): the identification of digital priorities for palliative care research using a modified Delphi method": Interquartile Range to be used to guide the Level of Agreement for Delphi

### Appendix: Interquartile Range to be used to guide the Level of Agreement for Delphi responses

#### Level of Agreement

- “Very high agreement” – median 5; percentage agreement ≥80%; IQR 0
- “High agreement” – median 4/5; percentage agreement ≥80%; IQR 1
- “Moderate agreement” – median ≤4; percentage agreement 60–79%; IQR 1
- “Low agreement” – median <4; percentage agreement <60%; IQR >1
