## Supplementary material for "Technology in Palliative Care (TIP): the identification of digital priorities for palliative care research using a modified Delphi method": Level of agreement for priority areas both Delphi rounds

**Appendix: Level of agreement for each 'priority area' following both Delphi rounds**

| Q | Description | Round 1 | | | | Round 2 | | | |
|---|---|---|---|---|---|---|---|---|---|
| **Q** | **Description** | **Median Rating (n=66)** | **IQR** | **% Agreement (4/5 on scale)** | **Level of Agreement** | **Median Rating (n=50)** | **IQR** | **% Agreement (4/5 on scale)** | **Level of Agreement** |
| **1.1** | Development of Electronic Health Records (EHR) and systems based approaches to collection and utilisation of Big Data | 4 | 1 | 76% | Moderate | 1 | 4 | 80% | High |
| **1.2** | Patient-Generated Health Data (PGHD) – what to collect and how: harnessing and using 'physical function' data via sensor-based technologies (edited) | 4 | 1 | 60% | Moderate | 1 | 4 | 62% | Moderate |
| **1.3** | Governance in the use of 'Big Data' – regulation and responsibility | 5 | 1 | 83% | High | 1 | 5 | 80% | High |
| **1.4** | Cybersecurity and ransomware – Packaging, capturing and transferring data and keeping it safe | 5 | 1 | 80% | High | 1 | 5 | 82% | High |
| **1.5** | Ethical Challenges in Big Data Health Research: implications for informed consent and participation | 4 | 1 | 78% | Moderate | 1 | 5 | 82% | High |
| **2.1** | Smart Threads – early development experimental stages but with positive results as a potential diagnostic or monitoring technology, or as a mechanism for displaying information | 3 | 1 | 45% | Low | 1 | 3 | 42% | Low |
| **2.2** | Physical Activity Change Detection (PACD) | 3 | 1 | 45% | Low | 1 | 3 | 36% | Low |
| **2.3** | Reducing burdensome interventions for patients: optimising monitoring and recording of physiological signals | 4 | 1 | 82% | High | 2 | 4 | 75% | Low |

| | | | | | | | | | |
|---|---|---|---|---|---|---|---|---|---|
| **2.4** | Wearable Health Trackers | 3 | 1 | 44% | Low | 1 | 3 | 36% | Low |
| **2.5** | Wearable fabrics for the detection of stimuli (physical functioning e.g. temperature, motion, strain, activity monitoring) | 3 | 2 | 36% | Low | 2 | 3 | 27% | Low |
| **2.6** | Body area network (BAN) technology | 3 | 2 | 30% | Low | 2 | 3 | 27% | Low |
| **2.7** | Wearable technology – privacy and ethical considerations | 4 | 2 | 67% | Low | 2 | 4 | 69% | Low |
| **3.1** | Use of mobile devices to gather patient reported symptom outcomes (PRO's)/Patient Performance Status | 4 | 1 | 81% | High | 1 | 4 | 89% | High |
| **3.2** | App design: Clinical input – Safety, efficacy, accuracy and assessment of risk | 4 | 1 | 76% | Moderate | 1 | 4 | 85% | High |
| **3.3** | Portable Hospital-level screening/diagnostics in the home | 4 | 2 | 73% | Low | 2 | 4 | 73% | Low |
| **4.1** | Telehealth/eHealth to support patients and their families in their own homes | 5 | 1 | 92% | High | 1 | 5 | 89% | High |
| **4.2** | Telehealth/eHealth for electronic patient-reported outcomes (ePRO) | 5 | 1 | 83% | High | 1 | 4 | 84% | High |
| **5.1** | Virtual reality (VR): Distraction therapy to ameliorate symptoms – pain, distress, anxiety | 4 | 2 | 73% | Low | 2 | 4 | 73% | Low |
| **5.2** | VR: grief and bereavement following the death of a patient | 4 | 2 | 50% | Low | 1 | 4 | 55% | Low |
| **5.3** | VR: for education and training needs | 4 | 1 | 80% | High | 1 | 4 | 82% | High |
| **6.1** | Artificial intelligence (AI) and Machine Learning (ML) to improve outcomes for individuals: Natural Language Processing (NLP) and systems-based approaches for prediction and 'screening' | 4 | 1 | 62% | Moderate | 1 | 4 | 56% | Low |
| **6.2** | Role of Big Data and AI/ML for Population Health Management – population level data | 4 | 1 | 62% | Moderate | 1 | 4 | 58% | Low |
| **6.3** | AI/ML – automation of human processes: ethical and moral issues | 4 | 2 | 58% | Low | 2 | 4 | 69% | Low |
| **7.1** | Robotics – for assistance and daily living | 4 | 1 | 50% | Low | 1 | 4 | 56% | Low |

| | | | | | | | | | |
|---|---|---|---|---|---|---|---|---|---|
| **7.2** | Robotics – for companionship/social inclusion | 3 | 2 | 40% | Low | 2 | 3 | 33% | Low |
| **7.3** | Robotics – optimisation of surgery | 4 | 2 | 64% | Low | 2 | 4 | 75% | Low |
| **7.4** | Robotics – Education | 4 | 1 | 53% | Low | 1 | 4 | 58% | Low |
| **8.1** | Smart Home Sensors – detect changes in health condition/physical function | 4 | 2 | 64% | Low | 1 | 4 | 65% | Moderate |
| **8.2** | Smart Home Sensors – Alert systems and monitoring – home security and controls | 4 | 2 | 64% | Low | 2 | 4 | 65% | Low |
| **8.3** | Smart Cities – Built Environment and Big Data | 3 | 1 | 41% | Low | 1 | 3 | 47% | Low |
| **9.1** | Genome profiling and Personalised Medicine | 4 | 2 | 66% | Low | 2 | 4 | 62% | Low |
| **9.2** | Genetic editing and biomarker technology for earlier disease detection and possible disease prevention | 4 | 2 | 68% | Low | 2 | 4 | 58% | Low |
